# The causal role of male pubertal timing for the development of externalizing and internalizing traits: results from Mendelian randomization studies

**DOI:** 10.1101/2024.05.30.24308257

**Authors:** Lars Dinkelbach, Triinu Peters, Corinna Grasemann, Anke Hinney, Raphael Hirtz

**Affiliations:** Department of Pediatrics III, University Hospital Essen, University of Duisburg-Essen, Essen, Germany; Institute of Sex- and Gender-sensitive Medicine, University Hospital Essen, University of Duisburg-Essen, Essen, Germany; Section of Molecular Genetics in Mental Disorders, University Hospital Essen, University of Duisburg-Essen, Essen, Germany; Center for Translational Neuro- and Behavioral Sciences, University Hospital Essen, University of Duisburg-Essen, Essen, Germany; Department of Pediatrics, Division of Rare Diseases and CeSER, St. Josef-Hospital, Ruhr-University Bochum, Bochum, Germany; Center for Child and Adolescent Medicine, Helios University Hospital Wuppertal, Witten/Herdecke University, Wuppertal, Germany

**Keywords:** Male puberty timing, adolescence, mental health, depression

## Abstract

Preexisting epidemiological studies suggest that early pubertal development in males is associated with externalizing (e.g., conduct problems, risky behavior, and aggression) and internalizing (e.g., depression and anxiety) traits and disorders. However, due to problems inherent to observational studies, especially of reverse causation and residual confounding, it remains unclear whether these associations are causal. Mendelian randomization (MR) studies take advantage of the random allocation of genes at conception and can establish causal relationships. In the current study, N=76 independent genetic variants for male puberty timing (MPT) were derived from a large genome-wide association study (GWAS) on 205,354 participants and used as an instrumental variable in MR studies on 17 externalizing and internalizing traits and psychopathologies utilizing outcome GWAS with 16,400 to 1,045,957 participants. In these MR studies, earlier MPT was significantly associated with higher scores for the overarching phenotype of ‘Externalizing Traits’ (beta=-0.03, 95%-CI [-0.06, -0.01]). However, this effect was likely driven by an earlier age at first sex (beta=-0.17, 95%-CI [-0.21, - 0.13]), without evidence for an effect on further externalizing phenotypes. Regarding internalizing phenotypes, earlier MPT was associated with higher levels of the ‘Depressed Affect’ subdomain of neuroticism (beta=-0.04, 95%-CI [-0.07, -0.01]). Late MPT was related to higher scores of internalizing traits in early life (beta=0.04, 95%-CI [0.01, 0.08]). In conclusion, this MR study supports a causal effect of MPT on specific traits and behaviors. However, no evidence for an effect of MPT on long-term clinical outcomes (depression, anxiety disorders, alcohol dependency, cannabis abuse) was found.

## Background

Mental health disorders and substance abuse have an enormous impact on overall disease burden, ranking among the top five contributors to disability-adjusted life years among all disease entities [1]. Interestingly, the disease burden attributable to mental disorders peaks during adolescence and young adulthood, with the highest burden in males aged 15-to 19-years [1].

Consistent with this observation, early pubertal timing has been suggested as a risk factor for externalizing (e.g., oppositional defiant disorder, antisocial behavior, risky sexual behavior, and substance abuse) and internalizing traits and disorders (e.g., depression or anxiety) in several cross-sectional epidemiological and clinical studies with small but consistent effects in both males and females [2]. However, these associations are subject to numerous confounders, including the social microenvironment (i.e., embedding in familial, peer, and school systems [3, 4], the macroenvironment (i.e., the neighborhood income) [3, 5], ethnicity [6], and family interaction patterns [7, 8] – most of which are difficult to control or correct for in observational studies.

Mendelian randomization (MR) studies can overcome limitations inherent to observational studies, such as reverse causation or residual confounding [9, 10]. In brief, MR studies take advantage of the random allocation of genetic variants at conception. If a genetic variant is associated with an exposure (e.g., puberty timing), this genetic variant can serve as an unconfounded predictor to analyze the effect of the exposure on the outcome of interest (e.g., psychopathologies) under certain assumptions [9, 10].

Several MR studies analyzed the effects of pubertal timing on psychopathologies in females and discovered an influence of early age at menarche on depression [11] and substance abuse [12]. In males, one MR study analyzed the effect of pubertal timing on substance addiction and found no effect on tobacco, alcohol, or cannabis addiction [12]. However, conclusions from this MR analysis are limited since both the exposure and outcome associations were drawn from a single sample, the UK Biobank, violating the non-sample overlap assumption of two-sample MR studies [13]. In addition, this study relied on a Genome-Wide Association Study (GWAS) assessing pubertal timing by single measures of physical maturation (such as late vs. early voice break and late vs. early facial hair growth) rather than a combined measure of male pubertal timing [12].

In males, a recent meta-analysis of GWAS based on 205,354 European males combined different indicators of pubertal timing in boys (e.g., the timing of voice break and facial hair growth) to study the genetic associations of male puberty timing (MPT) in general [14]. Thus, this large-scale GWAS is less dependent on a specific assessment of MPT and provides a more reliable tool to investigate the causal effect of male pubertal timing on psychopathologies. In a previous MR study, this GWAS was utilized to analyze the effect of MPT on depression but did not find robust evidence for a relationship between both [15]. However, further psychopathologies in males have not been addressed by MR studies so far, despite reasonable evidence for extensive associations of MPT with externalizing and internalizing traits and disorders in epidemiological studies [2].

### Objective

Here, we aim to utilize multiple MR studies based on a current large GWAS to analyze whether male pubertal timing has a causal effect on the development of 1.) externalizing traits, extraversion, antisocial behavior, and aggression, 2.) risky behavior, including general risk tolerance, smoking, and risky sexual behavior (number of sexual partners, age at first sex) 3.) substance abuse, i.e., alcohol and cannabis abuse, 4.) internalizing traits and neuroticism, and 5.) internalizing psychiatric entities such as depression and anxiety disorders.

## Methods

### Study design and assumptions

To address the above-mentioned research questions, 17 two-sample MR studies were conducted. In MR studies, genetic variants, usually single nucleotide polymorphisms (SNPs), are used as instrumental variables to analyze the effect of the genetically predicted exposure variable (here, MPT) on the outcome variable (here, externalizing and internalizing traits and psychopathologies). SNPs are valid instrumental variables if the three main assumptions of MR studies are met: (a.) the relevance criterion, meaning that the genetic variant has to be strongly related to the exposure, (b.) the exchangeability criterion, meaning that the genetic variants are not related to the outcome via other pathways than the exposure of interest, and (c.) the exclusion restriction criterion, meaning that the genetic variants are not directly related to the outcome [9]. In two-sample MR studies, the genotype-exposure associations and genotype-outcome associations are derived from two distinct GWAS analyses. Consequently, the two-sample MR approach relies on two additional assumptions: (e.) non-overlapping exposure and outcome GWAS as sample overlap can result in a bias of unknown direction and extent [13] and (f.) the assumption that both the exposure and outcome GWAS are derived from the same population [16].

Of the MR assumptions, only the relevance criterion (a.) can be tested directly [17]. This assumption was ensured by calculating F-statistics for each genetic variant. SNPs with F-statistics < 10 are considered weak instruments [9]. For MPT, only genome wide significant (p < 5x10^-8^) SNPs were utilized as instrumental variable [14], all surpassing the threshold for strong instruments (median F-statistic was 43.38, range 29.92 to 231.07). Violations of the second and third (b. and c.) assumptions arise when causal pathways between genetic variants and the outcome do not involve the exposure phenotype (i.e., the genetic variants are causally linked to the outcome either directly or via another phenotype), typically stemming from pleiotropy. Pleiotropy denotes a scenario where a genetic variant is linked to two or more seemingly unrelated phenotypes. To detect or correct for violations of the second and third (b. and c.) assumption, the following steps (detailed below) were taken: i.) identification and exclusion of pleiotropic outliers, ii.) calculating a set of so-called robust methods that are less susceptible to or correct for different kinds of pleiotropy [18], and iii.) correct for sources of pleiotropy by multivariable MR analyses [19]. Concerning step i.), the MR-PRESSO framework was used to identify horizontally pleiotropic outliers [20] and the identified pleiotropic SNPs were removed from further analysis. Concerning step ii.), the following set of robust methods were calculated after the exclusion of outliers: Weighted median-and mode-based estimates [21, 22], MR Egger [23] and MR-RAPS [24]. Details on these methods, how and to which extent they can correct for or are robust against pleiotropy, and their respective assumptions are given in Supplemental Methods 1. Concerning step iii.), increased body weight has consistently been associated with earlier puberty in boys [25, 26]. In addition, body weight is associated with internalizing and externalizing problems [27], suggesting it may serve as a pleiotropic pathway, which is supported by a previous MR study [15]. To analyze whether our results were affected by body weight-related pleiotropy, effect estimates of MPT on psychological outcomes were calculated, adjusting for BMI (IVW BMIcorr). This adjustment was made through multivariable MR analyses (MVMR). For MVMR, a GWAS on BMI in 374,756 males was used [28].

Despite efforts to identify outcome GWAS without sample overlap, the current study included several GWAS for relevant outcomes that used data from the UK Biobank, also contributing to the exposure GWAS addressing male puberty timing. Thus, to assess the impact of a violation of the ‘non-overlap assumption’ (e.), beta weights and standard errors for voice break (VB) were derived from 23andMe participants (N = 55,871; [14]) and used as an alternative instrumental variable, after exclusion of weak instruments (SNPs with F <10 for the phenotype VB), to calculate an effect estimate without sample overlap (IVW VB).

The primary endpoint for all MR analyses was the effect estimate, as calculated by the inverse variance weighted (IVW) method after excluding pleiotropic outliers as identified by the MR-PRESSO framework. To analyze whether the exclusion of outliers by MR-PRESSO biased the results, the raw IVW-effect estimate without outlier exclusion was calculated (MR-PRESSO raw) as a sensitivity analysis. To correct for multiple comparisons, the p-values of primary endpoints were corrected by the false discovery rate (FDR) [29]. Scatter plots (visualizing the effect of each SNP on male puberty timing and the outcome of interest), funnel plots (drawing the MR estimate of each SNP against the inverse of its standard error as a measure of its precision), and forest plots (illustrating the MR estimate for each SNP individually) were drawn to visually screen for violations of MR assumptions or distortion of effects by single genetic variants [18]. Leave-one-out analyses were conducted to assess whether the results were driven by single variants. The CheckSumStats package was used to confirm the correct annotation of the effect allele and the European population of the summary statistics utilized for the current MR analyses [30]. Details on the software packages used can be found in the Supplemental Methods. Figure 1 gives an overview of the study design.

**Figure 1.**
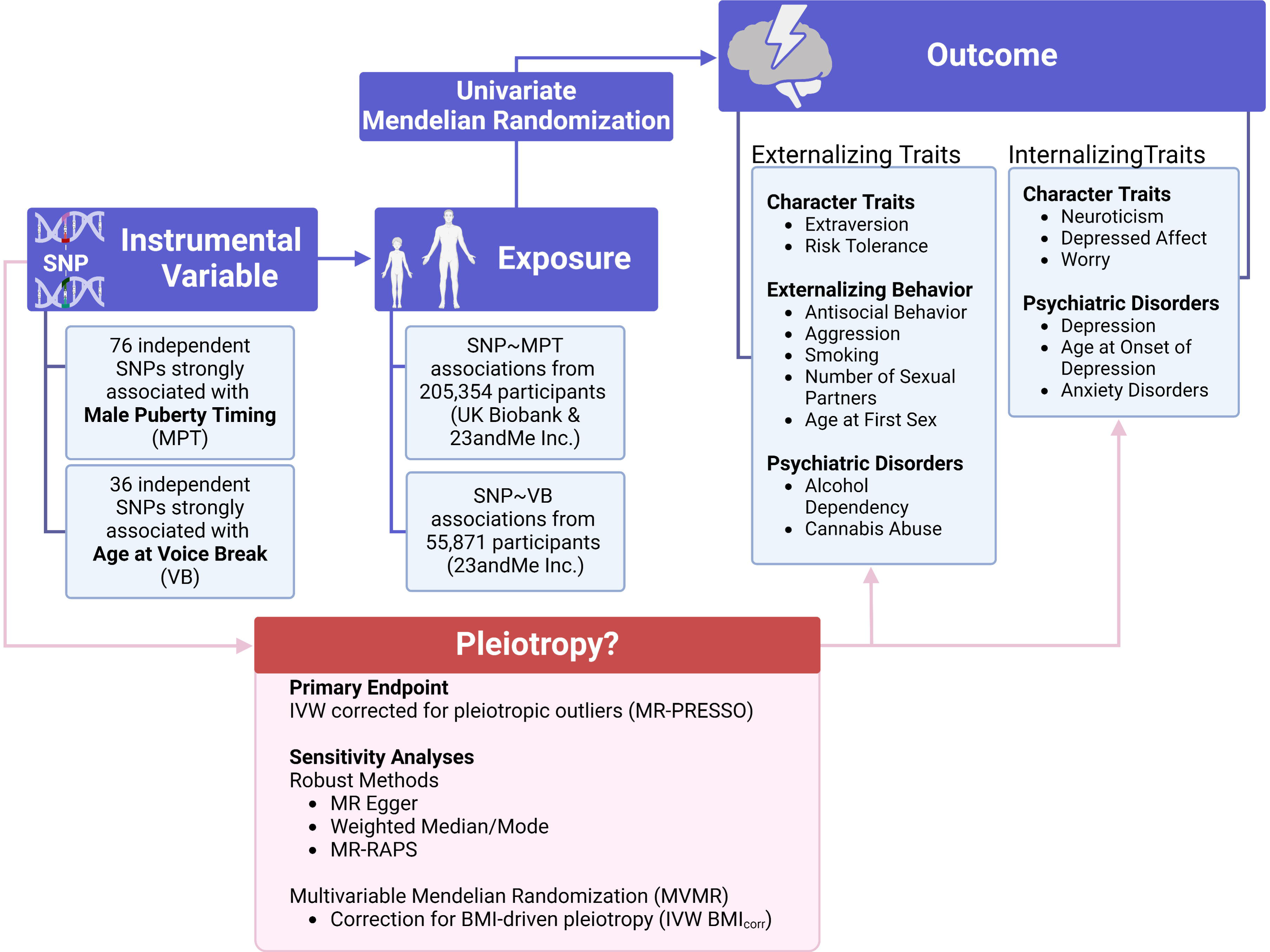
Illustration of the analytic approach and outcome variables assessed. The phenotypes ‘Antisocial Behavior’ and ‘Early Life Aggression’ as well as the overarching phenotypes ‘Externalizing Traits’ and ‘Early Life Internalizing Traits’ (the latter two are not shown for clarity) were based on meta-analytic GWAS pooling underlying traits, disorders, and behaviors [32-35, 44]. The character trait phenotypes ‘Extraversion’, ‘Risk Tolerance’, and ‘Neuroticism’ were based on cross-sectional measures with personality inventories (or based on a single item in the case of ‘Risk Tolerance’) in adulthood [36-38]. ‘Depressed Affect’ and ‘Worry’ represent genetically identified subclusters of ‘Neuroticism’ and thus were based on a subset of items of the ‘Neuroticism’ scale [37, 50]. The phenotype ‘Ever Smoker’ was based on the self-description of ever initiating smoking (i.e., smoking once or twice did not count as an ‘Ever Smoker’) [38]. For the ‘Number of Sexual Partners’ GWAS’ participants were asked about the number of sexual partners they had in their lifetime [38]. The phenotypes ‘Alcohol Dependency’, ‘Cannabis Abuse’, ‘Depression’, and ‘Anxiety Disorders’ were clinically defined based on the lifetime diagnosis of the respective psychiatric disorder using relevant diagnostic systems such as the DSM-IV or ICD-10 [39-42, 54]. The phenotypes ‘Age at First Sex’ and ‘Age at Onset of Depression’ were based on the recollected age at first sexual intercourse/age at onset of depressive symptoms or diagnosis [31, 43]. BMI=body mass index; IVW=inverse-variance weighted; SNP=single-nucleotide polymorphism

### Study population

Table 1 gives an overview of the GWAS and sample sizes utilized in the current study. The selection of outcome GWAS was motivated by i.) the sample size and novelty of the sample, ii.) the availability of summary statistics, and iii.) the potential sample overlap between exposure and outcome GWAS. However, the latter criterion was not always met due to the excessive use of consortia data to achieve large sample sizes. All participants of the GWAS utilized in the current study were of white European ancestry, thus presumably fulfilling the ‘same population assumption’ (f.). The focus on this population was driven by the higher availability of data rather than scientific interest. Only the outcome ‘Age at First Sex’ was based on a male-specific GWAS [31]. Due to the lack of large sex-specific GWAS, all other outcome GWAS were based on sex-combined GWAS. However, all sex-combined GWAS included sex as covariates (in some GWAS, no details on included covariates were provided). The phenotypes ‘Early Life Internalizing Traits’ and ‘Early Life Aggression’ were based on cross-sectional and longitudinal assessments in children and adolescents, leading to 251,152 observations of 64,561 subjects (for internalizing traits) and to 328,935 observations of 87,485 subjects (for aggression) included in the GWAS [32, 33]. About half of the observations in these two GWAS were examined in adolescents or late childhood (11-18 years) [32]. The remaining GWAS were mainly conducted in adults [31-44]. Supplemental Methods 2 and Supplemental Table S1 give a detailed description of how the respective phenotypes were obtained, the SNP-based heritability of each outcome GWAS, and further sample characteristics.

**Table 1.**
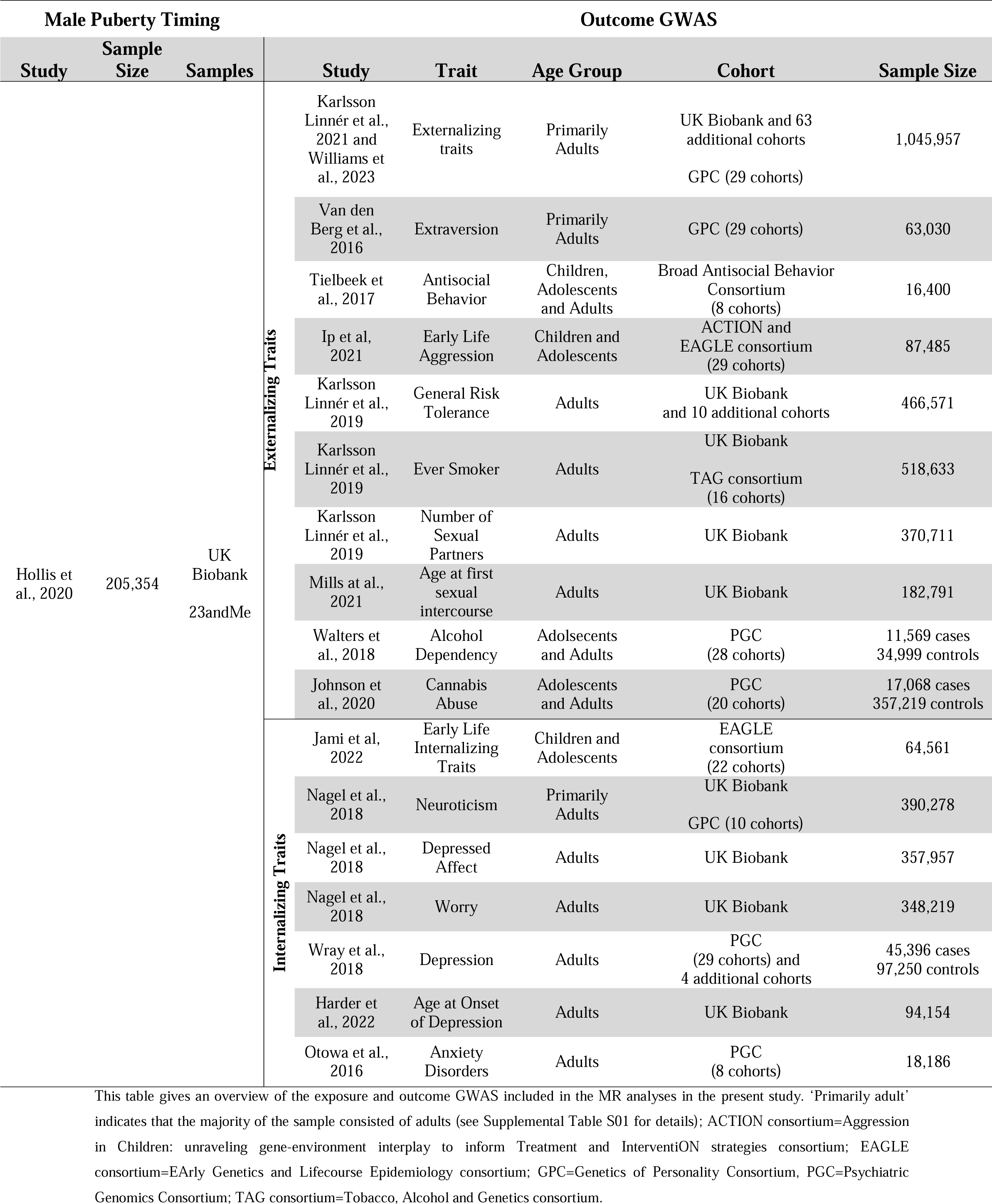
Overview of the exposure and outcome GWAS included in the MR analyses.

### Instrumental variable

For MPT, the GWAS by Hollis, et al. [14] based on 205,354 men of European ancestry was used. This GWAS comprised data from the UK Biobank and 23andMe in which MPT was operationalized as recalled age at voice break (in the 23andMe cohort) or, for UK Biobank participants, having a relatively (compared to peers) early/late (or about average) onset of voice break and onset of facial hair grow [14]. These five phenotypes (age at voice break, early/late voice break, early/late facial hair growth) were combined in a meta-analysis to perform a GWAS on the underlying trait ‘Male Puberty Timing’. In total, 76 independent (defined by distance of at least 1 MB) genome-wide significant (p<5x10^-8^) target SNPs for MPT were identified and utilized as instrumental variables for the current MR analyses. These SNPs explained 2.2% of the variance in MPT. The median F-statistic was 43.38 (range 29.92 to 231.07).

To evaluate if using UK Biobank data led to phenotype-exposure-related sample overlap that confounded our MR results, we conducted sensitivity analyses with ‘Age at Voice Break’ data from 23andMe Inc., which included 55,871 participants [14]. Of the 76 target SNPs for MPT, 36 had F-statistics > 10 for ‘Age at Voice Break’ (VB) had and thus qualified as instrumental variable for VB. For VB, beta and standard errors of these 36 SNPs were derived from the subsample from 23andMe Inc. participants as reported by Hollis, et al. [14].

## Results

### The effect of male puberty timing on externalizing traits and disorders

For ‘Externalizing Traits’, 65 of the 76 target SNPs for MPT were available in the outcome GWAS by Williams, et al. [34] for ‘Externalizing Traits’. The MR-PRESSO global test for pleiotropy was significant (RSS_obs_=380.38, p<3x10^-4^), and seven outliers were identified by MR-PRESSO (see Supplemental Table S02 for details). Thus, the instrumental variable used for further analyses consisted of 58 SNPs. The IVW estimate revealed a negative effect of genetically predicted male puberty timing on externalizing traits, i.e., earlier pubertal timing was related to higher scores for externalizing traits (IVW, beta=-0.03, 95%-CI [-0.06, -0.01], see Figure 2). The Q-statistic indicated significant heterogeneity (based on IVW: df=57, Q-statistic=139.70, p=6.78x10^-9^; based on MR Egger: df=56, Q-statistic=131.76, p=4.8x10^-9^). Sensitivity analyses: The weighted median, MR-RAPS, the IVW estimate without exclusion of pleiotropic SNPs (MR-PRESSO raw), and the MR analysis without sample overlap (IVW VB) confirmed the significant results (beta from -0.05 to -0.03, p-values from 0.002 to 0.007, see Figure 3A for details). The weighted-mode method and the MVMR considering BMI-related pleiotropy (IVW BMIcorr) led to similar effect estimates but with wider confidence intervals and were, thus, non-significant (Weighted Mode, beta=-0.04, 95%-CI [-0.09, 0.01], p=0.099; IVW BMIcorr, beta=-0.02, 95%-CI [-0.05, 0.01], p=0.151).

**Figure 2.**
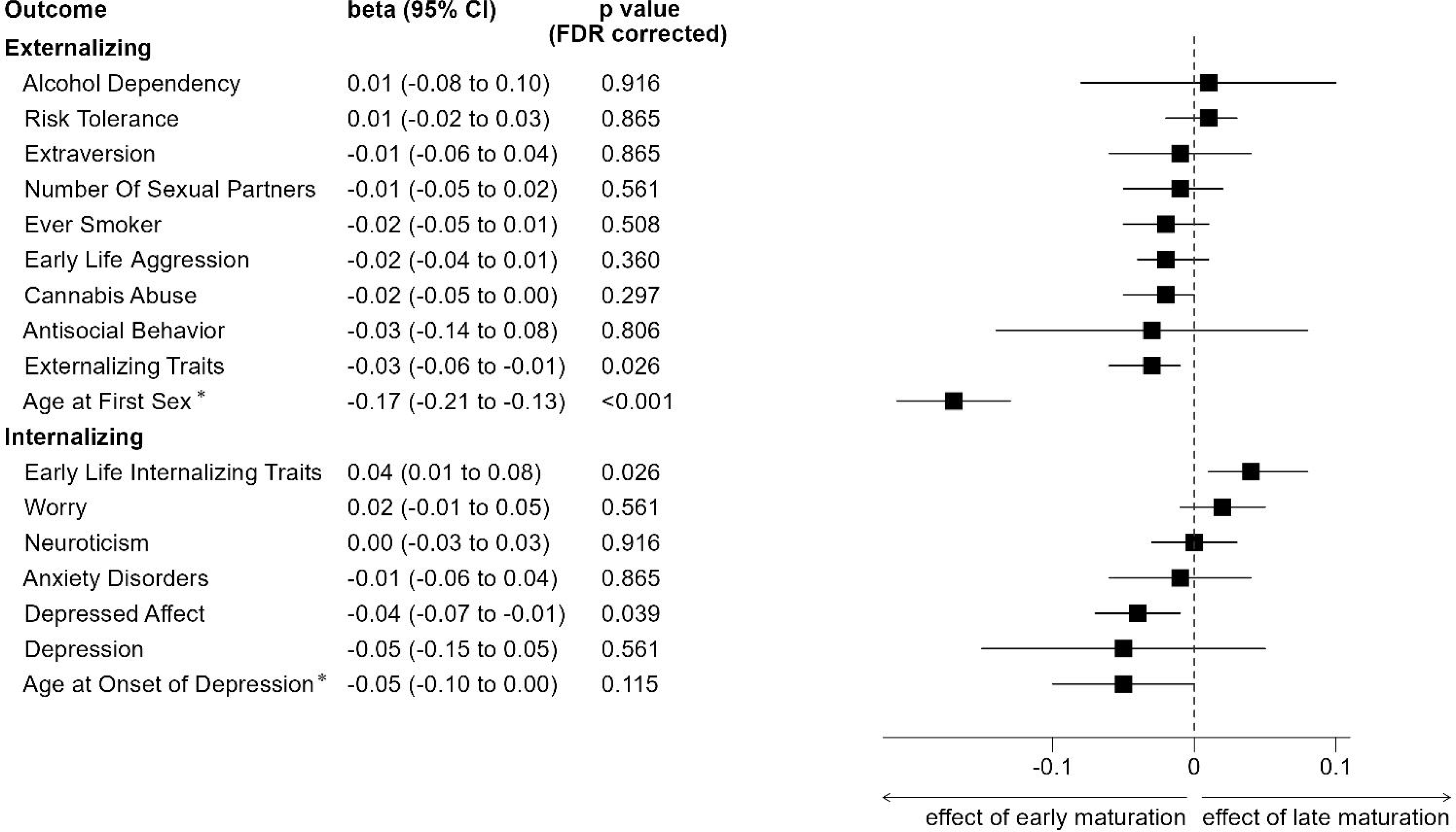
Results of the univariable Mendelian randomization (MR) analyses on the effect of male puberty timing on externalizing and internalizing traits and disorders. As primary endpoints, the inverse-variance weighted (IVW) effect estimates are illustrated after MR-PRESSO correction for pleiotropic outliers. * For reasons of comparability, ‘Age at First Sex’ and ‘Age at Onset of Depression’ inversely coded in the current MR analyses. Thus, negative effect estimates indicate a higher risk of developing psychopathology (for clinical endpoints), increased expression of a specific trait or behaviors, or a younger age at first sexual intercourse/depression onset with earlier male puberty timing. The outcome variables ‘Early Life Aggression’ and ‘Early Life Internalizing Traits’ are exclusively based on data from children and adolescents, whereas the other outcomes mainly involve adult cohorts (see Method Section and Supplemental Table S1 for a detailed description of cohorts and phenotypes). Results of the sensitivity analyses for FDR-corrected outcomes with a p-value < 0.05 are illustrated in Figure 3, while those for all other outcomes are given in Supplementary Figures S21-S33.

**Figure 3.**
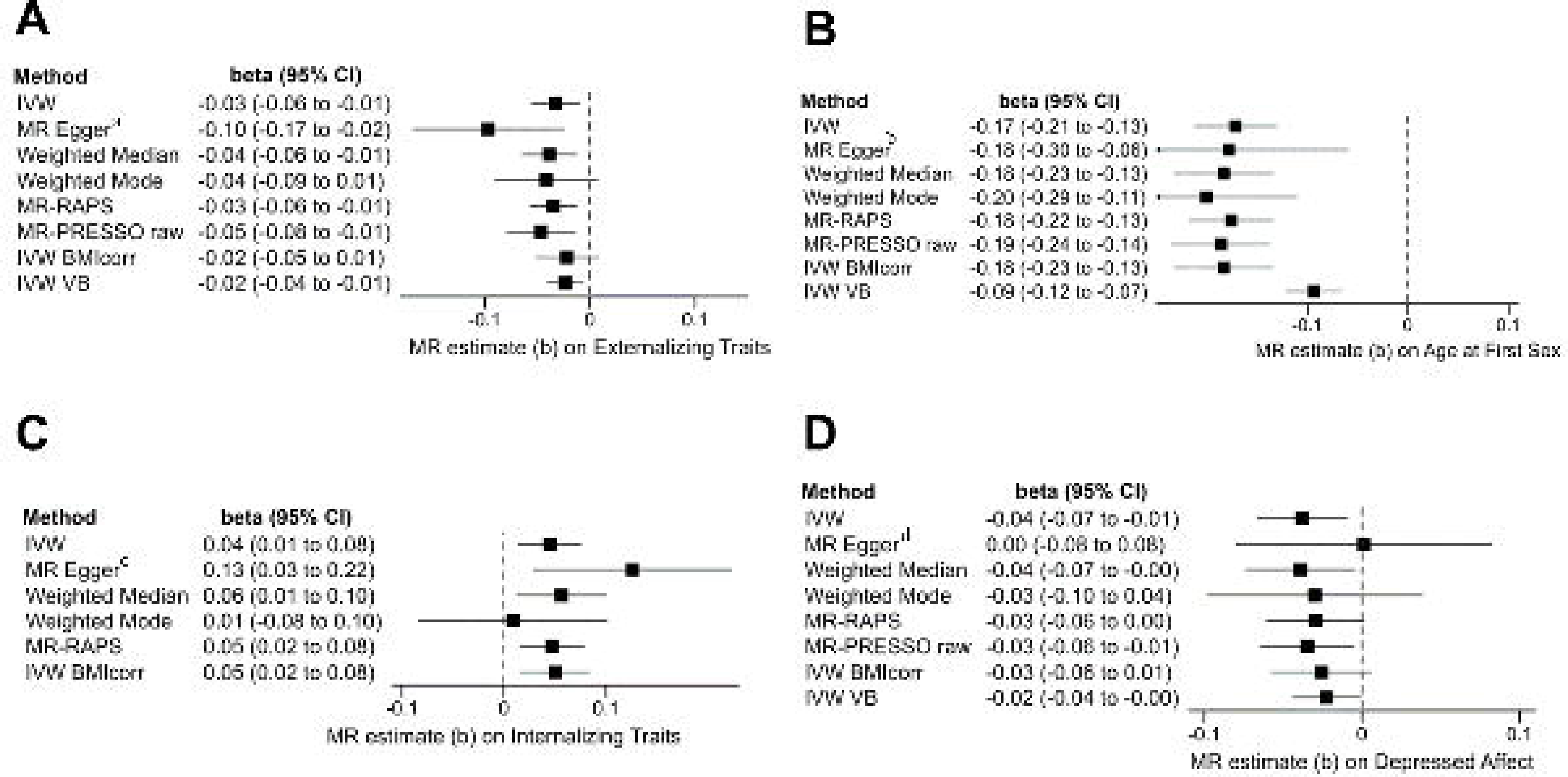
Results of the MR analyses of the effect of male puberty timing on the outcomes ‘Externalizing Traits’ (Fig. 3A), ‘Age at First Sex’ (Fig. 3B), ‘Early Life Internalizing Traits’ (Fig. 3C), and the ‘Depressed Affect’ subdomain of neuroticism (Fig 3D). ‘IVW’ represents the inverse-variance weighted (IVW) effect estimate after adjusting for pleiotropic outliers identified by MR-PRESSO. The ‘MR-PRESSO raw’ estimate represents the IVW estimate without excluding pleiotropic outliers. For the outcome ‘Early Life Internalizing Traits’ no pleiotropic outlier was detected, and thus, in this instance, the ‘MR-PRESSO raw’ effect estimate corresponds to the IVW estimate and therefore is not shown. The ‘IVW BMIcorr’ estimate represents the IVW effect estimate (after exclusion of pleiotropic outliers by MR-PRESSO) of male puberty timing on the specified outcome in an MVMR analysis considering BMI as an additional exposure. The ‘IVW VB’ effect estimate is the IVW effect estimate (after exclusion of pleiotropic outliers by MR-PRESSO) for the exposure ‘Age at Voice Break’ on the respective outcome. This exposure is exclusively based on 23andMe participants to ensure no exposure-outcome sample overlap, as none of the outcome GWAS datasets included 23andMe data. *a:* Eggers-intercept did not show evidence for significant directional pleiotropy (intercept=0.0018, se=0.0010, p=0.072). The *I*^2^-statistic was 0.762, indicating a possible violation of the NOME assumption. Therefore, the MR Egger result should be interpreted with caution. *b*: Eggers-intercept did not show evidence for significant directional pleiotropy (intercept=0.0002, se=0.0017, p=0.905). The *I*^2^-statistic was 0.777, again indicating a possible violation of the NOME assumption. *c*: Eggers-intercept did not show evidence for significant directional pleiotropy (intercept=-0.0002, se=0.0014, p=0.089). The *I*^2^-statistic was 0.785, indicating a possible violation of the NOME assumption. *d*: Eggers-intercept did not show evidence for significant directional pleiotropy (intercept=-0.0013, se=0.0012, p=0.314). The *I^2^*-statistic was 0.780, indicating a possible violation of the NOME assumption for the conduction of MR Egger.

For ‘Age at First Sex’, all the 76 target SNPs for male puberty timing were covered in the outcome GWAS by Mills, et al. [31]. The MR-PRESSO global test for pleiotropy was significant (RSS_obs_=241.57, p<3x10^-4^), and MR-PRESSO identified five outliers. The MR analysis revealed a significant effect of MPT on the age at first sexual intercourse; earlier MPT was associated with an earlier age at first sexual intercourse (IVW, beta=-0.17, 95%-CI [-0.21, -0.13], see Figure 2). The Q-statistic revealed significant heterogeneity (IVW: df=70, Q-statistic=138.83, p=1.9x10^-6^; MR Egger: df=69, Q-statistic=183.80, p=1.3x10^-6^). All sensitivity analyses, including those addressing heterogeneity, confirmed this effect (beta from -0.20 to -0.09; see Figure 3B for details).

For the remaining externalizing outcomes (‘Alcohol Dependency’, ‘Risk Tolerance’, ‘Extraversion’, ‘Cannabis Abuse’, ‘Number of Sexual Partners’, ‘Ever Smoker’, ‘Early Life Aggression’, and ‘Antisocial Behavior’), the IVW effect estimate remained non-significant (see Figure 2). For ‘Risk Tolerance’, ‘Number of Sexual Partners’, ‘Cannabis Abuse’ and ‘Ever Smoker’, single sensitivity analyses indicated significant effects, but these no longer appeared in the remaining or additionally performed sensitivity analyses (see Supplementary Figures S23, S25, S26, and S27 for details).

### The effect of male puberty timing on internalizing traits and disorders

71 of the 76 target SNPs for ‘Male Puberty Timing’ were available in the outcome GWAS by Jami, et al. [32] for ‘Early Life Internalizing Traits’. The MR-PRESSO global test for pleiotropy was not significant (RSS_obs_=84.24, p=0.169), and MR-PRESSO identified no outlier. The MR analysis revealed a significant effect of genetically predicted male puberty timing: later maturation leads to more internalizing traits in children and adolescents (IVW, beta=0.04, 95%-CI [0.01, 0.07], see Figure 2). The Q-statistic indicated no significant heterogeneity (IVW: df=70, Q-statistic=81.88, p=0.157; MR Egger: df=69, Q-statistic=78.51, p=0.203). The above finding was confirmed by all sensitivity analyses (beta from 0.04 to 0.13), except for the weighted mode method (beta=0.01, 95%-CI [-0.08, 0.10], see Figure 3C).

Concerning the “Depressed Affect” subdomain of neuroticism, 72 target SNPs were included in the outcome GWAS by Nagel, et al. [37]. The MR-PRESSO global test for pleiotropy was significant (RSSobs=140.03, p<3x10^-4^), and one outlier was identified, which was subsequently excluded from further analyses (see Supplemental Table S2 for details). The MR analysis revealed that earlier MPT is associated with an increased ‘Depressed Affect’ (IVW, beta=-0.04, 95%-CI [-0.07, -0.01], see Figure 2). The Q-statistic was significant, indicating heterogeneity (IVW: df=70, Q-statistic=123.84, p=7.6x10^-5^; MR Egger: df=69, Q-statistic=122.03, p=8.7x10^-5^). Sensitivity analyses: The weighted median method, the IVW effect estimate without exclusion of outliers (MR-PRESSO raw), and the analysis excluding sample overlap (IVW VB) led to significant negative effect estimates (beta from – 0.04 to -0.02). The weighted mode method, MR-RAPS, and the MVMR (IVW BMIcorr) method provided similar, but less precise (meaning wider confidence intervals) effect estimates with non-significant hypotheses testing (beta from – 0.03 to -0.02, see Figure 3D for details). In contrast, the MR Egger method provided an effect estimate close to zero and wide confidence intervals (beta=0.00, 95%-CI [-0.08, -0.08]). However, the MR Egger method might have been biased by a possible violation of the NOME assumption (*I^2^*=0.780, see Supplemental Methods 1 for details). The funnel plot illustrating the precision of each genetic variant and their MR effect estimate revealed one outlier with a strong negative effect estimate (see Supplementary Figure S14). However, even after excluding this outlier, leave-one-out analyses confirmed an effect of MPT on ‘Depressed Affect’ (see Supplementary Figure S16).

For the outcome ‘Age at Onset of Depression’, a significant effect of ‘Male Puberty Timing’ was observed (IVW, beta=-0.05, 95%-CI [-0.10, 0.0], p=0.034 without FDR-correction), suggesting that earlier maturation in males is linked to a younger age at onset of depression. However, after FDR correction, this effect must be considered non-significant according to conventional criteria (p=0.115, see Figure 2). Sensitivity analyses, including MR Egger, MR-RAPS, MR-PRESSO raw, and IVW VB, supported an association between MPT and ‘Age at Onset of Depression’. In contrast, the weighted median and weighted mode method yielded non-significant results with smaller (weighted median, beta=-0.02, 95%-CI [-0.09, 0.04],) and even positive (weighted mode, beta= 0.01, 95%-CI [-0.14, 0.17]) effect estimates, suggesting a potential bias of the IVW results by pleiotropy (see Supplementary Figure S21 for details). Leave-one-out analyses further underscored the instability of this finding, as the exclusion of individual SNPs rendered the association between MPT and ‘Age at Onset of Depression’ non-significant (see Supplementary Figure S20 for details).

For all other internalizing outcomes, MR analyses yielded non-significant results, see Figure 2). This pattern was confirmed across various sensitivity analyses (see Supplementary Figures S22-S33 for details).

## Discussion

Epidemiological studies indicate that an early onset of puberty is a risk factor for a broad spectrum of externalizing and internalizing behaviors and disorders. However, correlational relationships in epidemiological studies are subject to numerous confounders. Here, several MR studies were performed to provide evidence for a causal contribution of MPT to the development of character traits, behaviors, and psychopathologies.

### Externalizing Traits and Disorders

There was evidence that early puberal timing in males leads to an earlier initiation of sexual activity, i.e., a younger age at first sexual intercourse. This observation is consistent with a genetic correlation between the age of voice break and the age at first sexual intercourse [31] as well as increased risky sexual behaviors in early maturating boys in epidemiological studies [2]. Our analyses also revealed an association between an early onset of puberty in boys and the development of externalizing traits. The outcome ‘Externalizing Traits’ was operationalized via a meta-analysis that combined seven problematic behaviors and psychiatric entities (including smoking, cannabis and problematic alcohol use, risk tolerance as well as the number of sexual contacts, and the age at first sexual contact; see Supplemental Methods 2 and [35]). However, except for the age at first sexual intercourse, MR analyses exploring the relationship between MPT and specific entities (‘Alcohol Dependency’, ‘Cannabis Abuse’, ‘Ever Smoker’, ‘Risk Tolerance’, ‘Number of Sexual Partners’) by individual GWAS found no significant effects of MPT on these outcomes (see Figure 2). Therefore, the statistical relationship between MPT and the combined outcome ‘Externalizing Traits’ seems primarily driven by the effect of MPT on age at first sexual contact, and it remains unclear whether MPT has further influence on externalizing traits beyond this effect.

### Internalizing Traits and Disorders

To our surprise, we observed that genetically predicted *later* male puberty leads to more internalizing traits in children and adolescents (‘Early Life Internalizing Traits’; i.e., depressive symptoms, emotional problems, and anxiety). This observation contrasts with most previous epidemiological studies suggesting a link between *early* (but not late) onset of puberty and the development of internalizing problematic traits in adolescent boys [2]. Additionally, a meta-analysis of the effects of *late* puberty timing in males did not indicate an association with the occurrence of psychopathologies [2]. However, this conclusion rests on a limited number of studies, as noted by the authors of this meta-analysis [2]. Further, a recent large combined epidemiological and MR study including the most comprehensive set of confounders to date, found no evidence of a relationship between (either early or late) MPT and major depressive disorder (MDD) in males [15]. This finding suggests that previous studies may have been affected by confounding, which could also extend to other domains of internalizing traits. The ‘gendered deviation’ hypothesis postulates that those experiencing the most extreme deviations in puberty timing are at the highest risk for mental health problems [2, 45]. As girls generally mature earlier than boys, late maturing boys are particularly at risk of suffering psychosocial stress, as late maturing boys are not only the last of their sex, but also the last of their whole age group to go through puberty [2, 45]. So far, epidemiological studies suggested little evidence for this hypothesis [2]. In contrast, our observation of more internalizing symptoms with genetically predicted later maturation in boys would be consistent with this hypothesis. However, it must be noted that the outcome GWAS by Jami, et al. [32] relied on a heterogeneously defined phenotype of ‘Early Life Internalizing Traits’ that was assessed using multiple sources (e.g. parental, self or teacher ratings) and different measurement instruments, across a broad age range (boys and girls between 3 and 18 years). As individual-level data or age-/instrument-specific GWAS were not available to us, our analysis cannot disentangle whether the effect found here is age-specific (e.g., later MPT leading to more internalizing problems, early MPT leading to less internalizing problems, or both) or non-linear (e.g., extreme early or late puberty leading to more internalizing problems). Additionally, it remains unclear whether our finding is applicable to all measurement approaches included to define the outcome phenotype. Thus, the final interpretation of our results, and the extent to which confounding factors may account for discrepancies with existing epidemiological studies, must be determined by future, well-designed epidemiological studies that are able to analyze age-specific and non-linear effects and complement our approach.

### Long-term effects of MPT

Although numerous cross-sectional studies suggest a relationship between earlier onset of puberty and (clinical) endpoints (i.e., the development of alcohol dependency, cannabis abuse, major depression, or anxiety disorders), this was not shown by the MR analyses conducted in the present study. The clinical outcomes in the present study were assessed in adults considering acute or past diagnoses or symptoms, thereby serving as an estimate of the lifetime risk for these conditions. The ‘maturational disparity’ hypothesis postulates that early maturing children and adolescents are confronted with a mismatch between rapid physical changes and slowly evolving cognitive resources to cope with these changes [2, 4, 46] which is also supported on a neurophysiological level [47, 48]. However, whether this mismatch attenuates with increasing age, and thus, the long-term impact of early puberty on problematic behaviors still needs to be investigated. Recent longitudinal studies have shown that the effects of early puberty in males on antisocial behavior, depression, externalizing traits, and substance abuse decrease (and in some cases even reverse) with increasing age [25, 49]. Accordingly, the MR analyses conducted here also showed a relationship between an early onset of puberty and age at first sexual contact but not the total number of sexual partners in adulthood. Also, while there was a notable trend towards a younger age of onset of depressive disorders, this did not apply to the lifetime risk for the development of a major depressive disorder (MDD). Thus, these results indicate that mental health consequences associated with ‘maturational disparity’ are transient, possibly confined to adolescence and early adulthood. On a genetic level, a recent GWAS in 94,154 participants with depression found a significant, albeit not perfect, genetic correlation (r_g_ = -0.49) between age at onset of depression and lifetime MDD risk [43]. Similarly, strong but not perfect correlations were found between internalizing traits in children and adolescents and the lifetime risk for depression and anxiety disorders (r_g_ = 0.71 and 0.76; [32]). Thus, even though much of the genetic liability of psychiatric disorders is shared across the lifespan, their manifestation exhibits age-related specificity. Further GWAS focusing on specific age groups (e.g., in children, adolescents, and young adults) are needed to gain a deeper understanding of these complex relationships across the lifespan and could inform future MR studies.

The only lifetime trait affected by MPT was the ‘Depressed Affect’ subdomain of neuroticism, with early puberty timing in males leading to increased score for ‘Depressed Affect’. The ‘Depressed Affect’ subdomain is derived from four of twelve items of a neuroticism scale (for details, see Supplemental Methods 2) by clustering genetic correlations on a single-item level [50]. In contrast, the other subdomain of this scale, ‘Worry’, exhibited a non-significant effect estimate in the opposite direction in the present study. Consequently, our MR studies revealed a non-significant effect of MPT on the broad trait ‘Neuroticism’. Despite strong genetic correlations between ‘Depressed Affect’ and depressive symptoms as well as anxiety [50], no effect of MPT on these outcomes was seen here. Thus, further studies on the relevance of these subdomains of ‘Neuroticism’ are needed. However, it should be noted that the effect of MPT on ‘Depressed Affect’ was not consistently found in sensitivity analyses, potentially related to issues of pleiotropy.

### Limitations

Our study has several limitations. First, the exposure (MPT) was determined based on self-reported information on the timing of voice breakage and facial hair growth, which might be subject to recall bias. However, polygenic risk scores derived from the target SNPs for MPT significantly predicted longitudinal self-reported Tanner staging in 1,126 boys [14]. Second, GWAS for 16 of the 17 outcomes analyzed in the current study were based on sex-pooled effect estimates rather than sex-specific analyses, primarily because sex-specific GWAS were not available. However, current evidence suggests marginal sex differences in the autosomal genetic architecture of most traits, behaviors, and psychopathologies [51], justifying their use as outcomes in MR with a sex-specific exposure. Accordingly, this approach has been applied in various MR studies, including testosterone levels in men or age at menarche concerning MDD [11, 52]. Despite evidence of sex differences in the genetic liability for the trait ‘Antisocial Behavior’, we decided to use the sex-combined GWAS with 16,400 participants, as the male-stratified GWAS included only 7,772 participants and would therefore probably not have been sufficiently powered. Male-specific summary statistics with an adequate sample size (182,791) were only available for the outcome ‘Age at First Sex’. Third, the sample sizes of the outcome GWAS varied widely, leading to large differences in the power of the conducted MR studies. As most of the outcome GWAS were based on continuous outcomes and the power estimation of MR studies on continuous outcomes is dependent on further information, e.g. on the exposure-outcome association on the phenotypic level [53], which was not available to us, we decided against power calculations based on arbitrary assumptions. However, despite reasonable sample sizes in most of the studies used here, minor effects of MPT might have been missed. Fourth, two-sample MR studies as utilized here are based on linear models and therefore do not account for potential non-linear relationships.

### Conclusions

The MR analyses conducted here support a causal effect of MPT on the development of specific psychological traits and behaviors. Specifically, earlier MPT was associated with an earlier age at first sex, while later pubertal timing was associated with increased internalizing traits in children and adolescents. However, the effects of male puberty timing might be transient as no effects on adulthood extraversion or neuroticism or lifetime risk for psychiatric disorders were found. In line with this conclusion, there was a notable trend towards earlier age at the onset of depression with early maturation. However, this trend did not perpetuate to the lifetime risk of developing a major depressive disorder. To further explore the causal relationships between puberty timing and the onset of psychopathologies, particularly during the critical periods of late adolescence and early adulthood, additional age-and sex-specific GWAS followed by Mendelian randomization analyses are needed.

## Ethics

All GWAS utilized in the current MR studies were conducted after approval of the respective local ethics committee and obtained written informed consent of the participants or their legal representatives.

## Author contributions

LD, TP, and RH conceptualized the study. LD conducted the statistical analyses. TP and RH supervised the statistical analyses. LD wrote the first draft of the manuscript. TP, CG, AH, and RH critically revised the manuscript.

## Supporting information

Supplemental Methods and Results

## Data Availability

All data produced in the present study are available upon reasonable request to the authors.

## Abbreviations

BMI: body mass index
FDR: false discovery rate
GWAS: genome-wide association study
IVW: inverse-variance weighted
MDD: major depressive disorder
MPT: male puberty timing
MR: Mendelian randomization
MVMR: multivariable Mendelian randomization
SNP: single-nucleotide polymorphism

