## Supplemental Methods and Results for "The causal role of male pubertal timing for the development of externalizing and internalizing traits: results from Mendelian randomization studies"

### Overview

### Supplementary Methods 1 – Statistical methods to calculate effect estimates

### Supplementary Methods 2 – Outcome GWAS and phenotypes

### Supplementary Methods 3 – Software

### Supplementary Results 1 – Details on outcomes with primary endpoint p value < 0.05

Externalizing Traits

Supplementary Figure S1 – Scatter Plot - Externalizing Traits

Supplementary Figure S2 – Funnel Plot - Externalizing Traits

### Supplementary Figure S3 – Forest Plot - Externalizing Traits

### Supplementary Figure S4 – Leave-one-out analysis - Externalizing Traits

Age at First Sex

Supplementary Figure S5 – Scatter Plot – Age at First Sex

### Supplementary Figure S6 – Funnel Plot – Age at First Sex

### Supplementary Figure S7 – Forest Plot – Age at First Sex

### Supplementary Figure S8 – Leave-one-out analysis – Age at First Sex

Early Life Internalizing Traits

Supplementary Figure S9 – Scatter Plot – Early Life Internalizing Traits

### Supplementary Figure S10 – Funnel Plot – Early Life Internalizing Traits

### Supplementary Figure S11 – Forest Plot – Early Life Internalizing Traits

### Supplementary Figure S12 – Leave-one-out analysis – Early Life Internalizing Traits

Depressed Affect

Supplementary Figure S13 – Scatter Plot – Depressed Affect

### Supplementary Figure S14 – Funnel Plot – Depressed Affect

### Supplementary Figure S15 – Forest Plot – Depressed Affect

### Supplementary Figure S16 – Leave-one-out analysis – Depressed Affect

Age at Onset of Depression

Supplementary Figure S17 – Scatter Plot – Age at Onset of Depression

### Supplementary Figure S18 – Funnel Plot – Age at Onset of Depression

### Supplementary Figure S19 – Forest Plot – Age at Onset of Depression

### Supplementary Figure S20 – Leave-one-out analysis – Age at Onset of Depression

Supplementary Figure S21 – Sensitivity Analyses – Age at Onset of Depression

### Supplementary Results 2 – Results of outcomes with primary endpoint p value > 0.05

Supplementary Figure S22 – Sensitivity Analyses – Alcohol Dependency

Supplementary Figure S23 – Sensitivity Analyses – Risk Tolerance

Supplementary Figure S24 – Sensitivity Analyses – Extraversion

Supplementary Figure S25 – Sensitivity Analyses – Cannabis Abuse

Supplementary Figure S26 – Sensitivity Analyses – Number of Sexual Partners

Supplementary Figure S27 – Sensitivity Analyses – Ever Smoker

Supplementary Figure S28 – Sensitivity Analyses – Early Life Aggression

Supplementary Figure S29 – Sensitivity Analyses – Antisocial Behavior

Supplementary Figure S30 – Sensitivity Analyses – Worry

Supplementary Figure S31 – Sensitivity Analyses – Neuroticism

Supplementary Figure S32 – Sensitivity Analyses – Anxiety Disorders

Supplementary Figure S33 – Sensitivity Analyses – Depression

Supplementary References

### Supplementary Methods

**Methods 1 – Calculation of effect estimates**

First, the MR-PRESSO method was calculated [1]. This method is based on three parts a.) the MR-PRESSO global test, which detects whether overall horizontal pleiotropy is present by comparing the observed effect of each SNP with the estimated effect (obtained via calculation of the effect estimate without the respective SNP). The observed residual sum of squares (RSSobs) is then compared with a simulated expected distribution of the residual sum of squares (using a Gaussian distribution), and this comparison is used to measure total horizontal pleiotropy. b.) The MR-PRESSO outlier test, where the same approach is used to detect horizontal pleiotropy of single variants, and c.) the MR-PRESSO distortion test, which evaluates whether the effect estimates differ with and without the exclusion of pleiotropic outliers [1]. As the primary endpoint for all outcomes, the inverse-variance weight (IVW) effect estimate was calculated after excluding pleiotropic outliers [2]. The IVW effect estimate combines the effect estimate for the exposure on the outcome for each SNP included in the IV, similar to a fixed-effect meta-analysis [2]. As sensitivity analyses, the following set of so-called robust methods was calculated: Weighted median- and mode-based estimates [3, 4], MR Egger [5], and MR-RAPS [6]. Weighted median- and mode-based analyses are less powered but give valid estimates even in situations when just the majority (or plurality, for mode-based estimation) of instruments with similar causal effect estimates fulfill the MR assumption [3, 4]. MR Egger corrects for directional pleiotropy by the introduction of an intercept in the resulting regression model and thus allows all genetic variants to be invalid variants [5]. However, MR Egger estimates are again less powered and sensitive towards violations of the ‘Instrument Strength Independent of Direct Effect’ (InSIDE) assumption, which postulates that the causal effect of a variant on the outcome and the direct (pleiotropic) effect of a genetic variant are uncorrelated. In addition, MR Egger relies on the ‘no measurement error’ (NOME) assumption, assuming that the association between each genetic variant and the exposure is measured without error [7]. A violation of these assumptions and its effect on the validity of the MR Egger test can be estimated by the *I^2^* statistic [7]. To assess violations of the NOME assumption, the *I*^2^ statistics were calculated for each MR Egger analysis. An *I*² < 0.9 indicate a relative bias in the MR Egger estimate greater than 10%, thus this value was used as a cut-off for a relevant violation of the NOME assumption [7]. Lastly, an MR-RAPS estimate was calculated, considering systematic pleiotropy by modeling the pleiotropic effect of each genetic variant with a random-effects distribution [6].

**Methods 2 - Outcome GWAS and phenotypes**

In this section, the outcome GWAS as well as the methods to assess the respective phenotypes are described. An overview of the GWAS characteristics is available in Table 1, a more detailed description can be found in Supplementary Table S01.

For **externalizing traits**, the summary statistic of the GWAS by Williams, et al. [8] based on an analysis by Karlsson Linnér, et al. [9] was used. The GWAS by Karlsson Linnér, et al. [9] pooled data of case-control GWAS (each with N > 50.000) for seven externalizing behaviors and disorders: general risk tolerance, attention-deficit/hyperactivity disorder (ADHD), problematic alcohol use, lifetime cannabis use, lifetime smoking initiation, age at first sexual intercourse (reversely coded), and number of sexual partners. Based on these data, a latent multi-trait variable of externalizing traits was defined. In total, up to 1,373,240 participants from 65 cohorts were included. Reported heritability varied from 5.3% (for general risk tolerance) to 26% (for ADHD). The largest cohorts came from 23andMe (N_max_ 599,289) and the UK Biobank (N_max_ 403,349). As the company 23andMe Holding Co. restricts access to the data of their samples, the GWAS was rerun based on 1,045,957 participants, excluding the 23andMe sample. This reanalysis yielded similar factor loadings and genetic correlations [8]. For the current MR analysis, the later GWAS without 23andMe data was used for the outcome ‘Externalizing Traits’.

For the personality trait ‘**Extraversion**’, a GWAS from the Genetics of Personality Consortium by Van den Berg, et al. [10] was used. This GWAS was based on a meta-analysis of 63,030 subjects of 29 cohorts and derived a latent variable ‘Extraversion’ based on the response behavior on extraversion items of different personality scales (see Supplementary Table S01 for details). SNP-based heritability was 5.0% [10].

For **‘Antisocial Behavior’**, a GWAS covering eight samples with 16,400 participants (children, adolescents, and adults) of European ancestry was utilized [11]. The trait ‘Antisocial Behavior’ was based on a broad definition by the diagnosis of antisocial personality disorders, retrospective or current diagnosis of conduct disorders, screening tools for antisocial behavior, or scales for rule-breaking behavior [11]. SNP-based heritability was 5.2% [11].

For ‘**Early Life Aggression’**, a GWAS by the ACTION (Aggression in Children: unraveling gene-environment interplay to inform Treatment and InterventiON strategies) and EAGLE (EArly Genetics and Lifecourse Epidemiology) consortium, covering data from 87,485 children and adolescents (1.5 to 18 year old) from 29 cohorts, was utilized [12]. As participants were longitudinally assessed, the GWAS was based on 328,935 observations. The phenotype was defined by 29 measures (self-report, parental report, or teacher ratings) covering scales of aggressive behavior, conduct problems, or oppositional defiant disorder [12]. SNP-based heritability was 3.1% [12].

For the outcomes ‘**Risk Tolerance**’, ‘**Ever Smoker**’ and ‘**Number of Sexual Partners**’ GWAS by Karlsson Linnér, et al. [13] were utilized. General risk tolerance was measured via a single-question item asking participants for their general comfort in taking risks. The GWAS included 466,571 participants from the UK Biobank and ten smaller replication samples. SNP-based heritability for ‘Risk Tolerance’ was 4.5%. The GWAS for ‘Ever Smoker’ was based on 518,633 participants of the UK Biobank and cohorts of the Tobacco, Alcohol, and Genetics (TAG) consortium, which was based on the participants’ self-description of being a former or current smoker. SNP-based heritability was 10.9%. For ‘Number of Sexual Partners’, data from 370,711 participants of the UK Biobank was used. Participants were asked about the number of sexual partners they had in their lifetime. SNP-based heritability for this trait was 12.8%.

The GWAS for ‘**Age at First Sex**’ was based on 182,791 male participants of the UK Biobank who were asked at which age they had their first sexual intercourse [14]. SNP-based heritability varied with the participants' birth year between 15% and 23%. For reasons of comparability, the age at first sex was inversely coded for this MR analysis.

For ‘**Alcohol Dependency**’ and ‘**Cannabis Abuse**’ as outcome variables, GWAS of the Psychiatric Genomics Consortium (PGC) were used. The GWAS of cannabis use disorder use was based on 357,219 controls and 17,068 cases that either met diagnostic criteria (ICD-10 codes or DSM-III-R/DSM-IV/DSM-V diagnoses) for cannabis abuse and/or dependence [15]. SNP-based heritability was 6.7%-12.1% (depending on the estimated population prevalence of this phenotype) [15]. The GWAS for alcohol dependency was based on 11,569 cases and 34,999 controls of European ancestry with a SNP-based heritability of 9% [16]. The diagnosis of alcohol dependency was made by clinicians or semi-structured interviews according to DSM-IV (or DSM-III-R) criteria [16].

For ‘**Early Life Internalizing Traits**’, a GWAS based on 22 samples included in the EAGLE consortium [17] was utilized. All participants conducted longitudinal assessments of internalizing symptoms, which were assessed by self-ratings (19.7%) or proxy-rating (80.3%, most often by mothers) based on established instruments, of which the Strengths and Difficulties Questionnaire (SDQ) and the Achenbach System of Empirically Based Assessment (ASEBA) were most often used (38.2% and 36.7% of cases). In total, 251,152 observations of 64,641 children and adolescents (3-18 years) were included in the resulting GWAS. About half of the observations (48.4%) were conducted in adolescence or late childhood (11-18 years). SNP-based heritability was 1.7% [17].

The outcome ‘**Neuroticism**’ and its two subdomains ‘**Depressed Affect**’ and ‘**Worry**’ were derived from the study by Nagel, et al. [18]. The measurement of the personality trait neuroticism was based on two different questionnaires (each containing 12 items). In a previous study, the authors analyzed the genetical architecture of neuroticism on a single-item level and thus identified two genetically distinct subclusters, ‘Depressed Affect’ and ‘Worry’, each based on four items [19]. While the GWAS for ‘Neuroticism’ utilized data from 390,278 participants of the Genetics of Personality Consortium (GPC) and UK Biobank, the GWAS for the two subclusters used UK Biobank data only (and thus led to slightly smaller sample sizes, see Table 1). SNP-based heritability was 10.0%, 8.9%, and 9.0% for ‘Neuroticism’, ‘Depressed Affect’, and ‘Worry’, respectively [18].

For ‘**Depression**’, a GWAS from the PGC by Wray et al. (2018) was used. This GWAS comprised fewer participants than other existing GWAS [20]. However, summary statistics cleared for participants of the UK Biobank were published; thus, this GWAS provides a non-overlapping sample with the exposure GWAS. In this down-sampled GWAS, 45,396 cases with a lifetime history of a major depressive disorder (as identified according to DSM-III, DSM-IV, ICD-9 or ICD-10 criteria via structured clinical interviews, checklists, or review of medical records) and 97,250 controls from 33 study cohorts were included [21]. SNP-based heritability of the down-sampled GWAS was not reported.

The GWAS for ‘**Age at Onset of Depression**’ was based on self-reported age at diagnosis and age of onset of symptoms of major depression in 94,154 participants of the UK Biobank [22]. SNP-based heritability was 5.6%. For reasons of comparability, this outcome was inversely coded for this MR analysis.

For **‘Anxiety Disorders’**, a GWAS conducted by Otowa, et al. [23] on 18,186 participants (including 7,016 subjects with anxiety disorders) of eight contributing samples of the PGC was used. The authors calculated a quantitative factor score that captures different degrees of clinical severity across different anxiety disorders (including subclinical forms) and served as the phenotype of the resulting GWAS. SNP-based heritability was 10.6% [23].

*Multivariable MR analysis with BMI*

An MVMR analysis was conducted for each outcome with BMI as an additional exposure to account for pleiotropy due to differences in body weight. Therefore, the GWAS on BMI in males was used from 374,756 participants from 29 studies of the Genetic Investigation of ANthropometric Traits (GIANT) consortium and from the UK Biobank [24].

*Data modification prior to MR analyses*

For the outcome ‘Early Life Internalizing Traits’, beta values were calculated from z-scores and se (beta = z * se). For the outcome ‘Depression’, beta values were calculated from odds ratios (beta = ln(OR)). For the outcomes ‘Neuroticism’, ‘Cannabis Abuse’, ‘Alcohol Dependency’ and ‘Antisocial Behavior’ only z-scores were available in the reported summary statistics. Beta values and standard errors were calculated from z scores, sample size and minor allele frequency (MAF) with the formula provided by Zhu, et al. [25]. For ‘Cannabis Abuse’, ‘Alcohol Dependency’ and ‘Antisocial Behavior’ no MAFs were reported, therefore these were approximated by the MAFs as reported in the GWAS on depression from Wray, et al. [21]. The GWAS by Wray, et al. [21] as well as the GWAS for ‘Cannabis Abuse’, ‘Alcohol Dependency’ and ‘Antisocial Behavior’ were based on consortia data with a presumably large sample overlap. Thus, the MAFs as reported by Wray, et al. [21] provided a good approximation for the MAFs of the GWAS for ‘Cannabis Abuse’, ‘Alcohol Dependency’ and ‘Antisocial Behavior’.

**Methods 3 - Software**

The exclusion of pleiotropic outliers and the MR-PRESSO global test were conducted with MR-PRESSO (version 1.0, [1]). The harmonization of genetic variants and the calculation of the IVW, MR Egger, the weighted median, and weighted mode estimators were conducted using the TwoSampleMR package (version 0.5.9, [26]). For harmonization of palindromic SNPs, it was assumed that all alleles were presented on the forward strand as the effect allele frequency was not available in all outcome GWAS to infer the forward strand. The MR-RAPS estimate (considering overdispersion and a I^2^-loss-function) was calculated with the mr.raps package (version 0.2, [6]). For the calculation of *I*^2^, the MendelianRandomization package was used (version 0.9.0, [27]). The MVMR analyses were conducted using the MVMR extensions of the MR-PRESSO and the MendelianRandomization package [1, 27]. The CheckSumStats package was used to confirm the correct annotation of the effect allele and the European population of the summary statistics utilized for the current MR analysis (version 0.0.0.9, only possible if the effect allele frequency was specified in the respective GWAS, [28]). Graphics were built with Rforestploter (version 1.1.1). All analyses were conducted using R version 4.3.2. F-statistics were calculated by F=beta²/se².

**Supplementary Results**

**Supplementary Results 1 – Details on outcomes with primary endpoint p value < 0.05**

**Outcome: Externalizing Traits**

65 out of the 76 target SNPs of male puberty timing were available in the outcome GWAS by Williams, et al. [8] and Karlsson Linnér, et al. [9]. MR-PRESSO identified seven pleiotropic SNPs (rs10188334, rs112881196, rs12203592, rs1514177, rs2049045, rs35063026, rs7853970, for details see Supplemental Table S02). Thus, the instrumental variable consisted of 58 genetic variants. The exclusion of outliers by MR-PRESSO did not alter the results of the primary endpoint (see ‘MR-PRESSO raw’ IVW estimate in Figure 3A). The scatter plots, funnel plots, forest plots, and leave-one-out analyses did not provide any evidence of violations of the Mendelian Randomization (MR) assumptions, nor were the results influenced by individual variants.

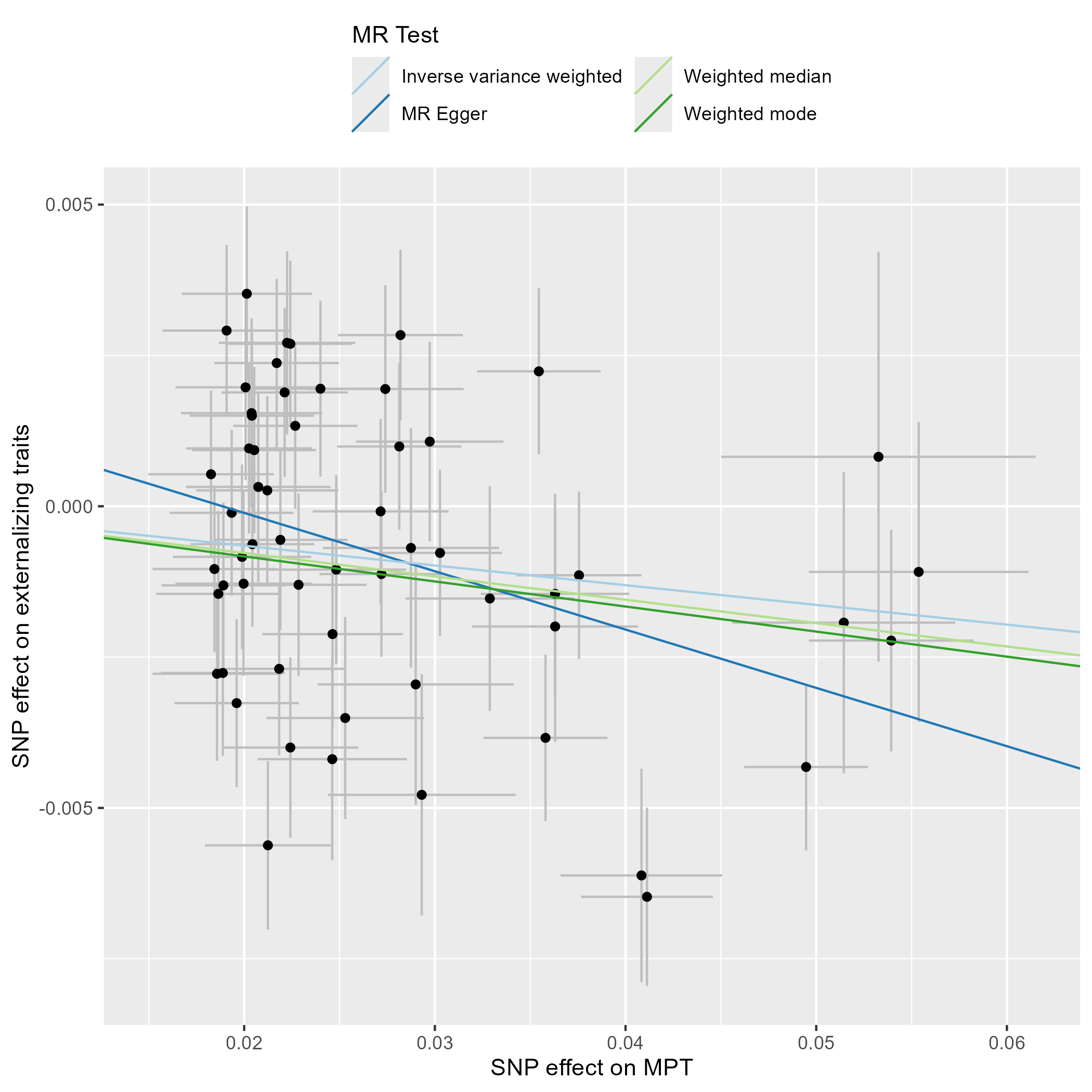

**Supplementary Figure S1 – Scatter Plot - Externalizing Traits.** This scatter plot depicts the effect estimates (beta, error bars represent the standard error) of each genetic variant included in the analysis on the exposure (male puberty timing [29]) and the outcome (externalizing traits, [9]).

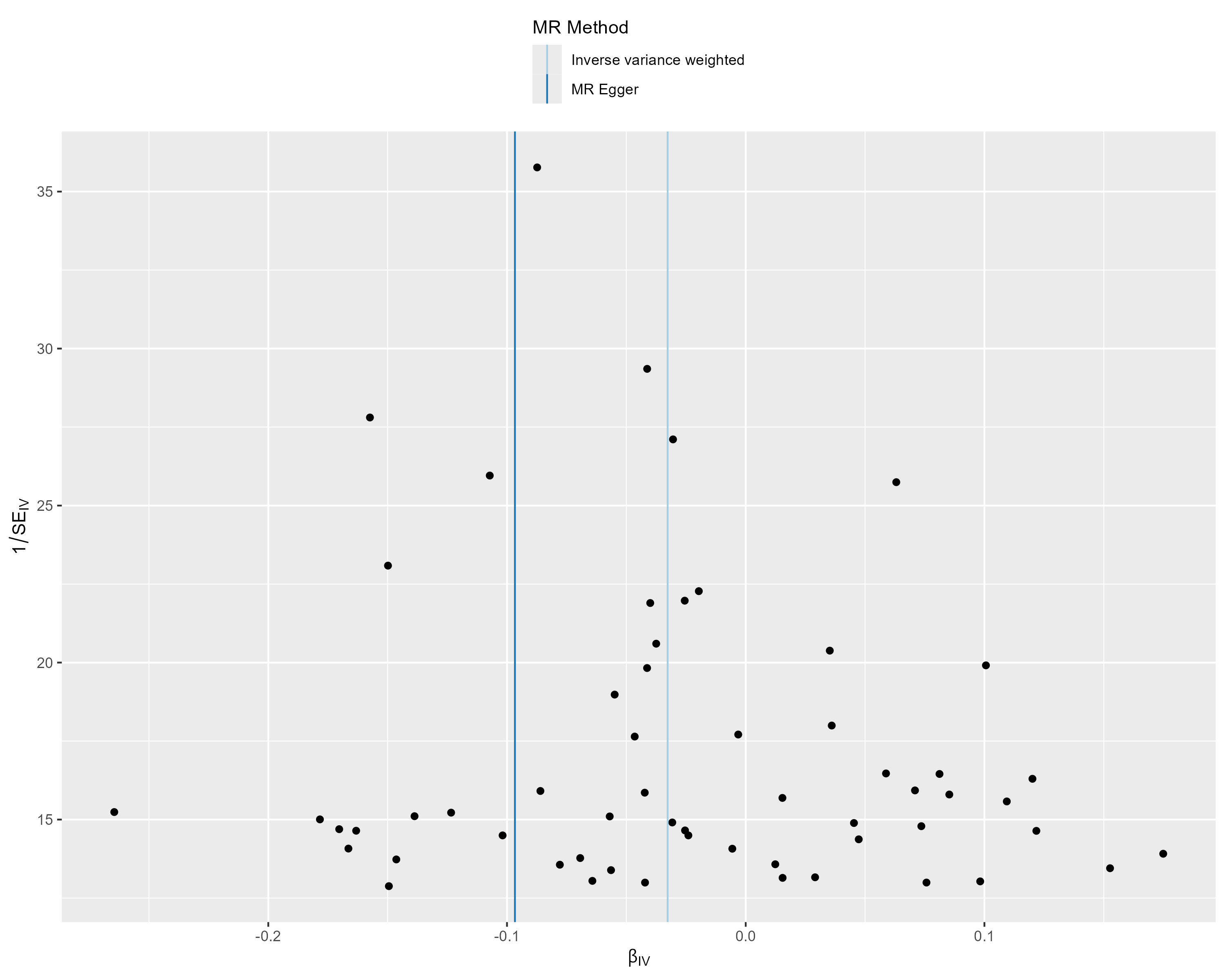

**Supplementary Figure S2 – Funnel Plot - Externalizing Traits.** This funnel plot illustrates the precision of each genetic variant (as measured by the inverse of the standard error (SE_IV_)) and their MR effect estimate (β_IV_).

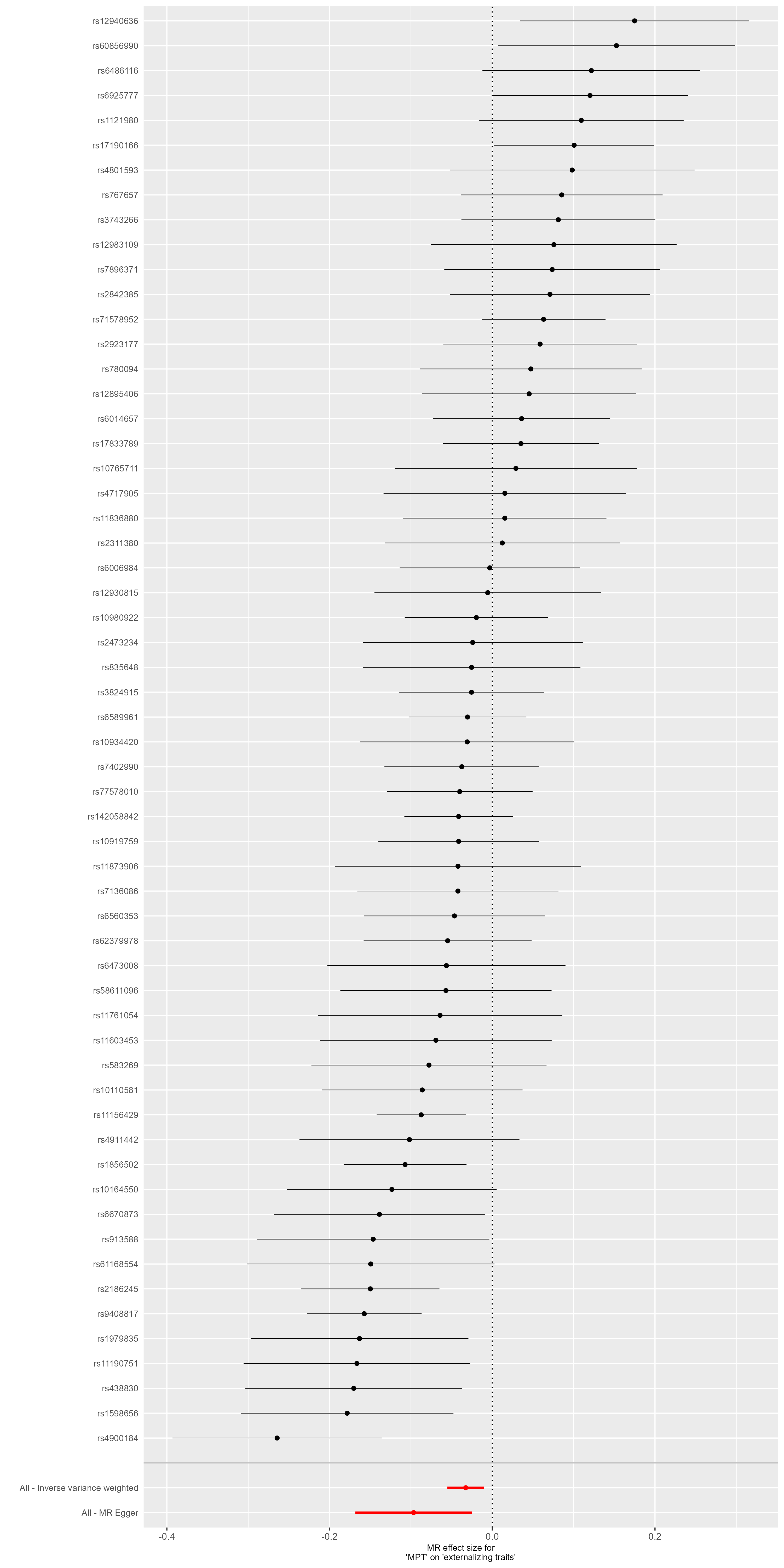

**Supplementary Figure S3 – Forest Plot - Externalizing Traits.** This forest plot illustrates the results of single-SNP MR analyses with the MR effect estimates for each SNP on the outcome (Externalizing Traits).

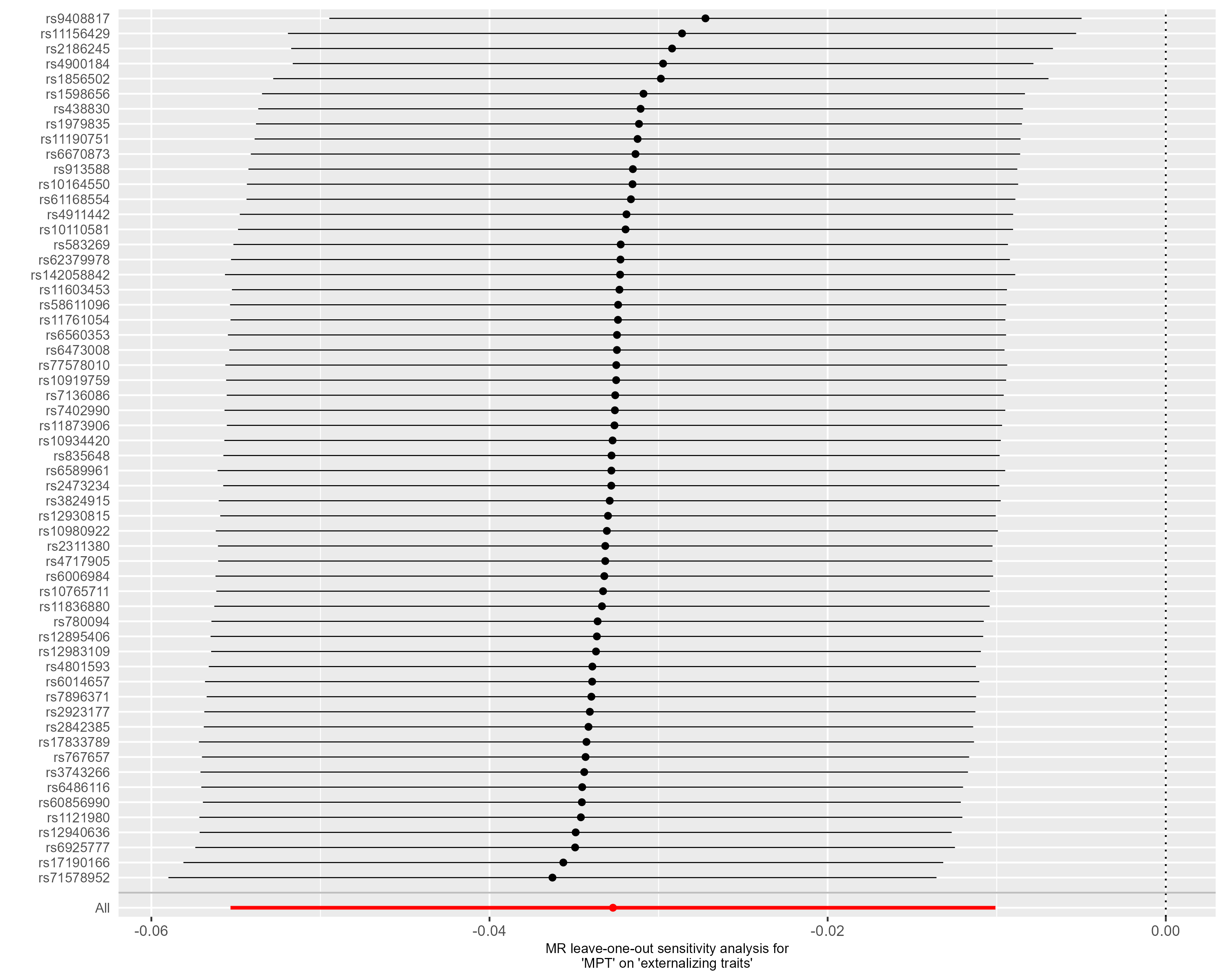

**Supplementary Figure S4 – Leave-one-out analysis - Externalizing Traits.** This forest plot illustrates the results of MR analyses (IVW) following the exclusion of individual genetic variants from the instrumental variable through leave-one-out analysis.

**Outcome: Age at First Sex**

All 76 target SNPs of male puberty timing were available in the outcome GWAS by Mills, et al. [14]. MR-PRESSO identified five pleiotropic SNPs (rs112881196, rs1514177, rs2049045, rs4900184, rs9408817), for details see Supplemental Table S2). Thus, the instrumental variable consisted of 71 genetic variants. The exclusion of outliers by MR-PRESSO did not alter the results of the primary endpoint (see ‘MR-PRESSO raw’ IVW estimate in Figure 3B). The scatter plots, funnel plots, forest plots, and leave-one-out analyses did not provide any evidence of violations of the Mendelian Randomization (MR) assumptions, nor were the results influenced by individual variants.

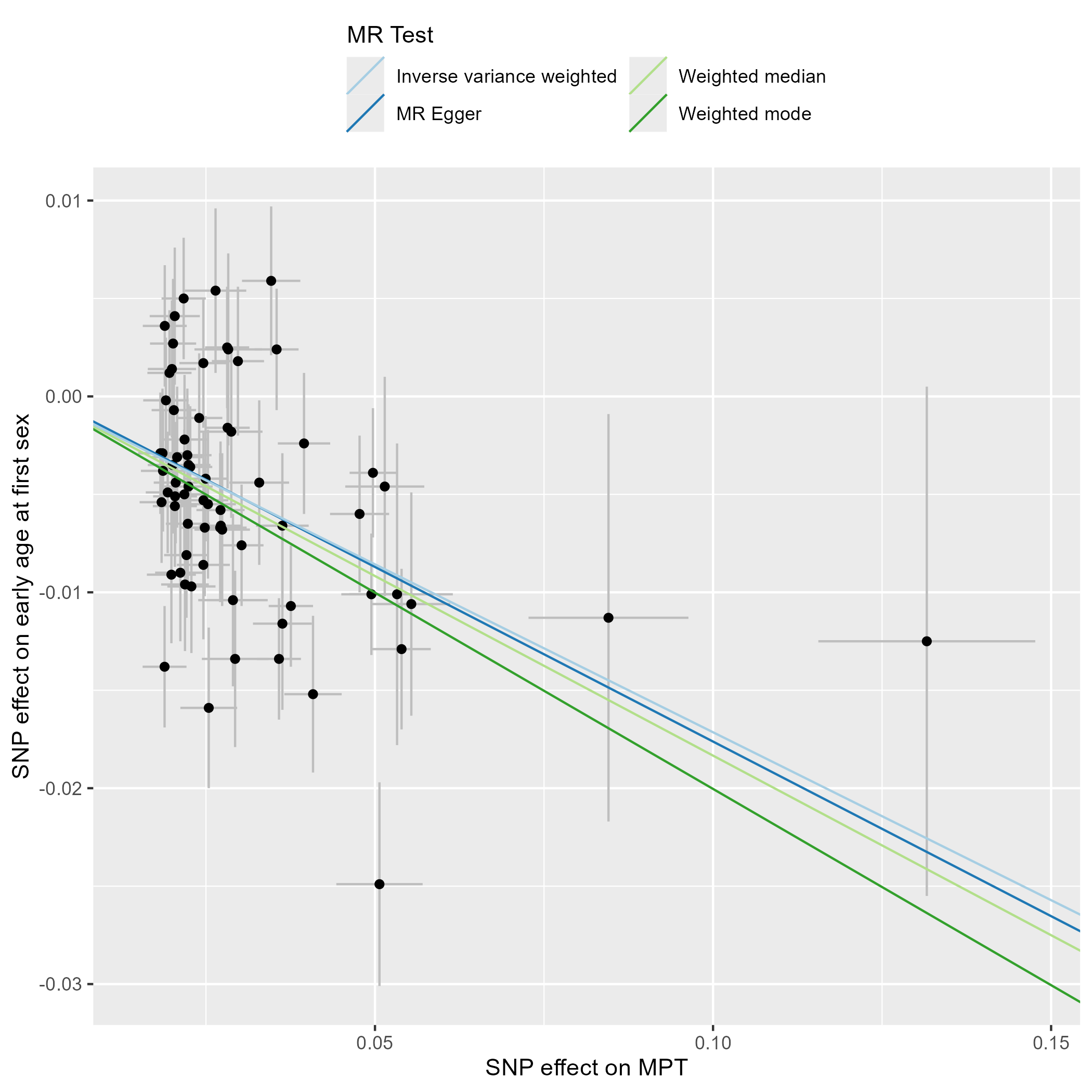

**Supplementary Figure S5 – Scatter Plot – Age at First Sex.** This scatter plot depicts the effect estimates (beta, error bars represent the standard error) of each genetic variant included in the analysis on the exposure (male puberty timing [29]) and the outcome (age at first sex, [14]).

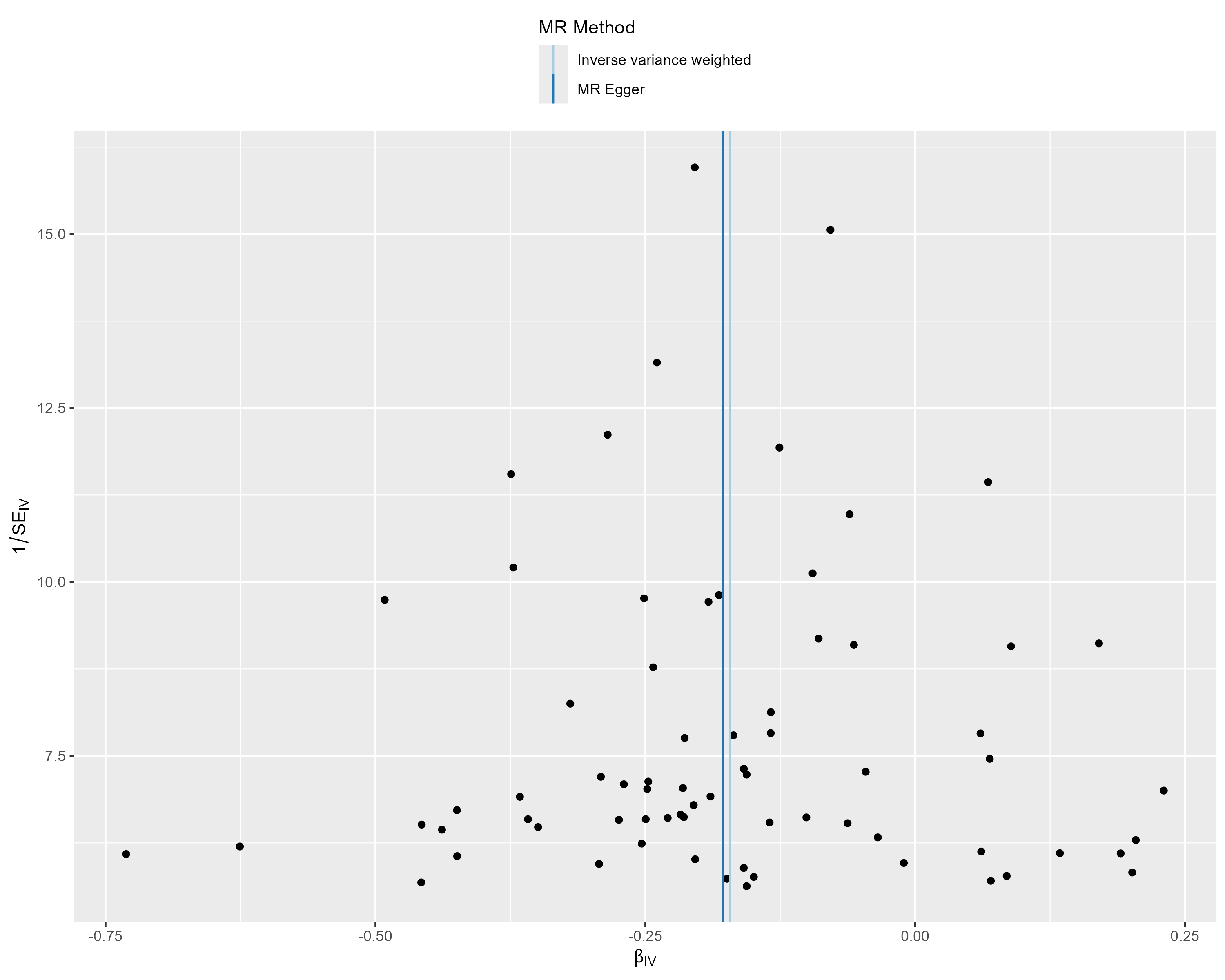

**Supplementary Figure S6 – Funnel Plot - Age at First Sex.** This funnel plot illustrates the precision of each genetic variant (as measured by the inverse of the standard error (SE_IV_)) and their MR effect estimate (β_IV_).

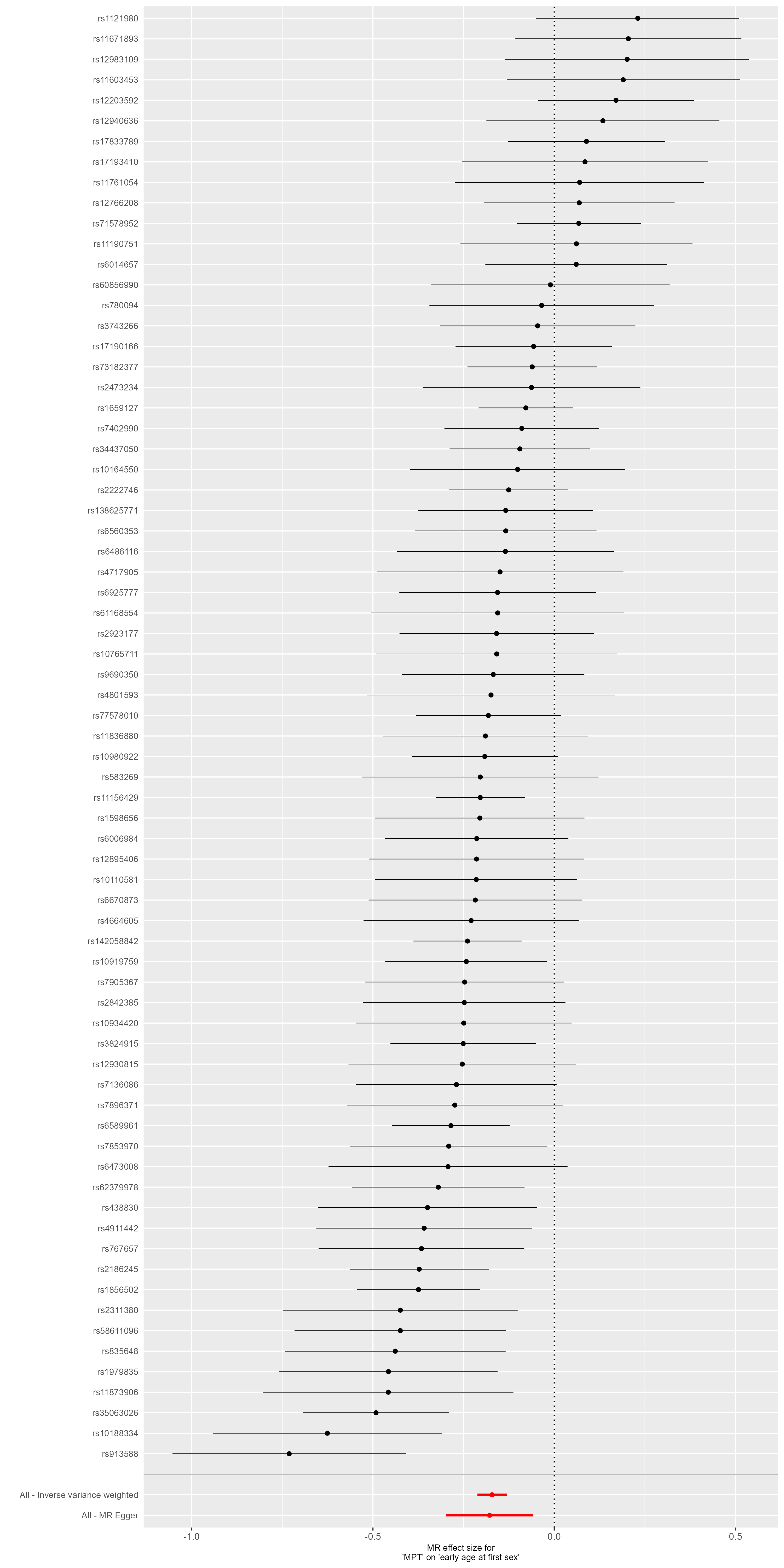

**Supplementary Figure S7 – Forest Plot - Age at First Sex.** This forest plot illustrates the results of single-SNP MR analyses with the MR effect estimates for each SNP on the outcome (Age at first sexual Intercourse, inversely coded). Negative effect estimates indicate that early maturation is related to younger age at first sexual intercourse.

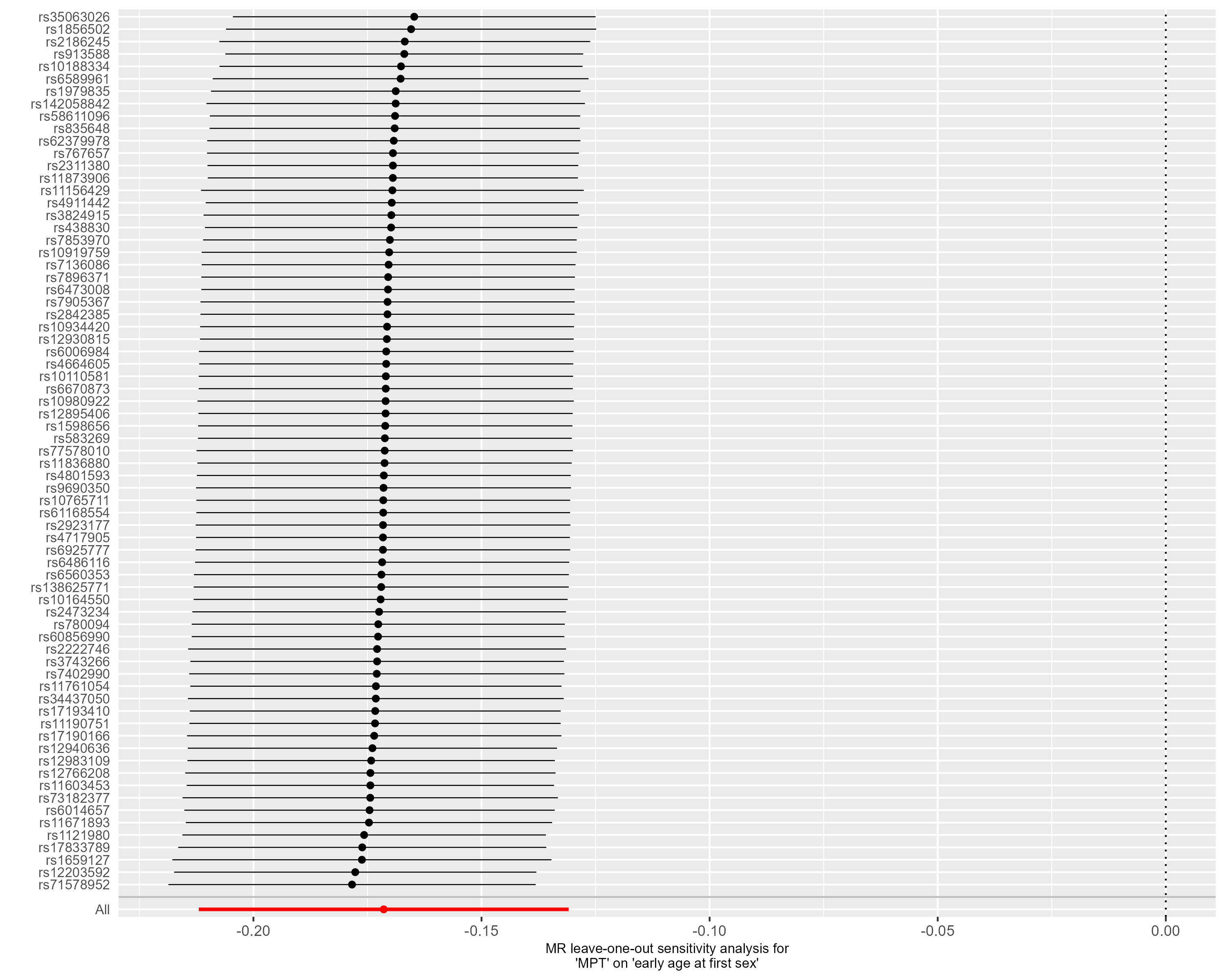

**Supplementary Figure S8 – Leave-one-out analysis - Age at First Sex.** This forest plot illustrates the results of MR analyses (IVW) following the exclusion of individual genetic variants from the instrumental variable through leave-one-out analysis.

**Outcome: Early Life Internalizing Traits**

71 of the 76 target SNPs of male puberty timing were available in the outcome GWAS by Jami, et al. [17]. No outliers were identified by MR-PRESSO. Thus, the instrumental variable consisted of 71 genetic variants. The scatter plots, funnel plots, forest plots, and leave-one-out analyses did not provide any evidence of violations of the Mendelian Randomization (MR) assumptions, nor were the results influenced by individual variants.

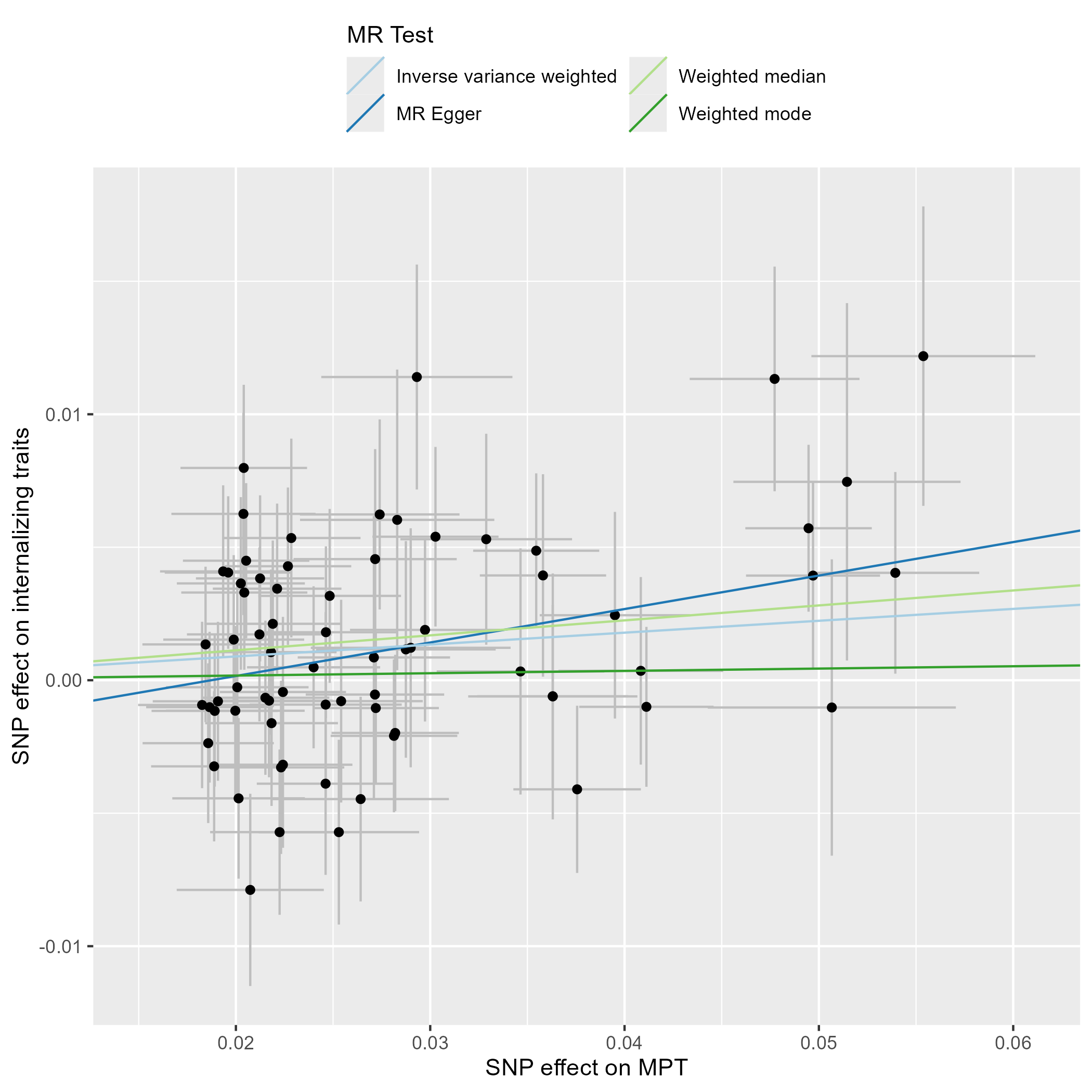

**Supplementary Figure S9 – Scatter Plot – Early Life Internalizing Traits.** This scatter plot depicts the effect estimates (beta, error bars represent the standard error) of each genetic variant included in the analysis on the exposure (male puberty timing [29]) and the outcome (internalizing traits in children and adolescents, [17]).

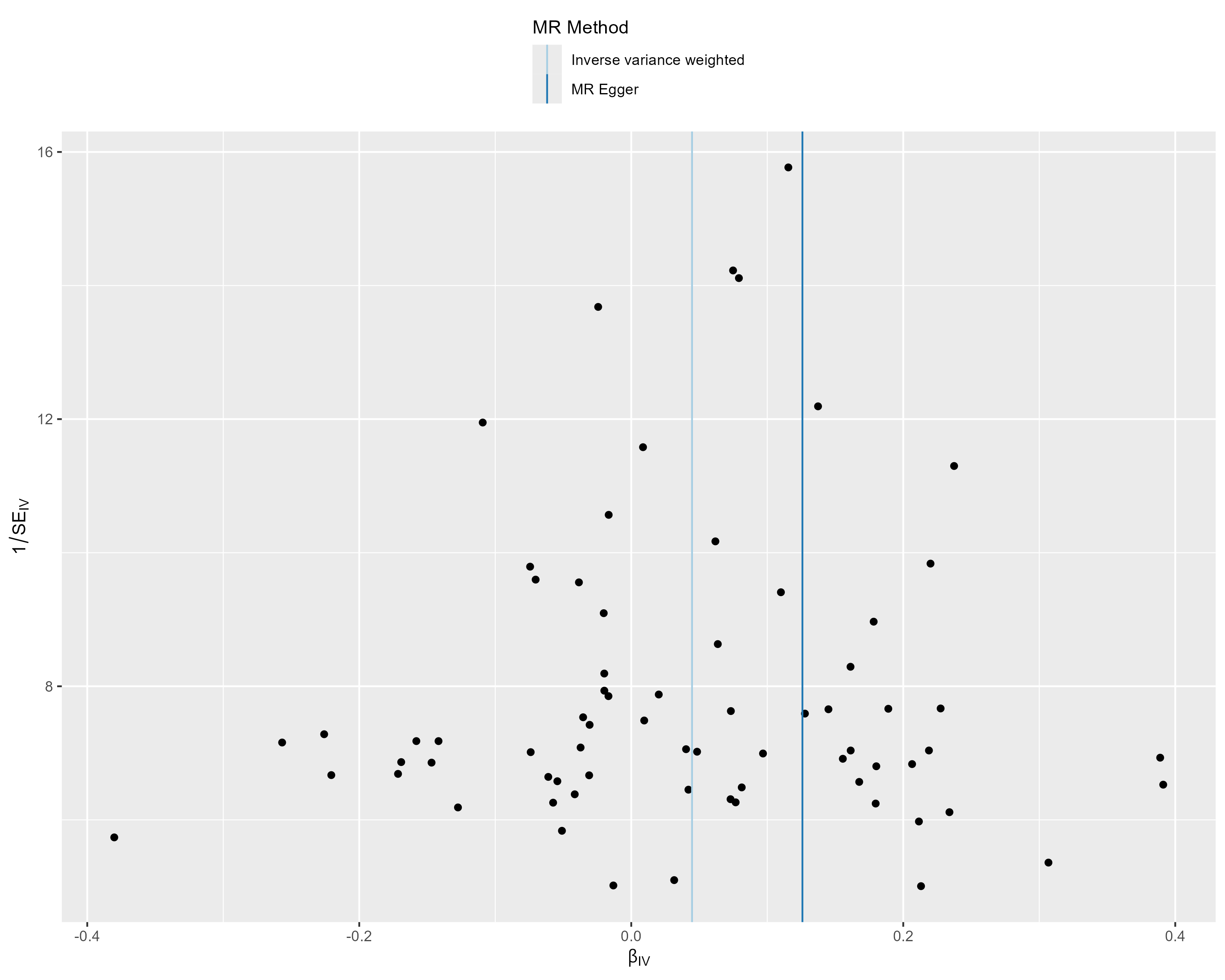

**Supplementary Figure S10 – Funnel Plot - Early Life Internalizing Traits.** This funnel plot illustrates the precision of each genetic variant (as measured by the inverse of the standard error (SE_IV_)) and their MR effect estimate (β_IV_).

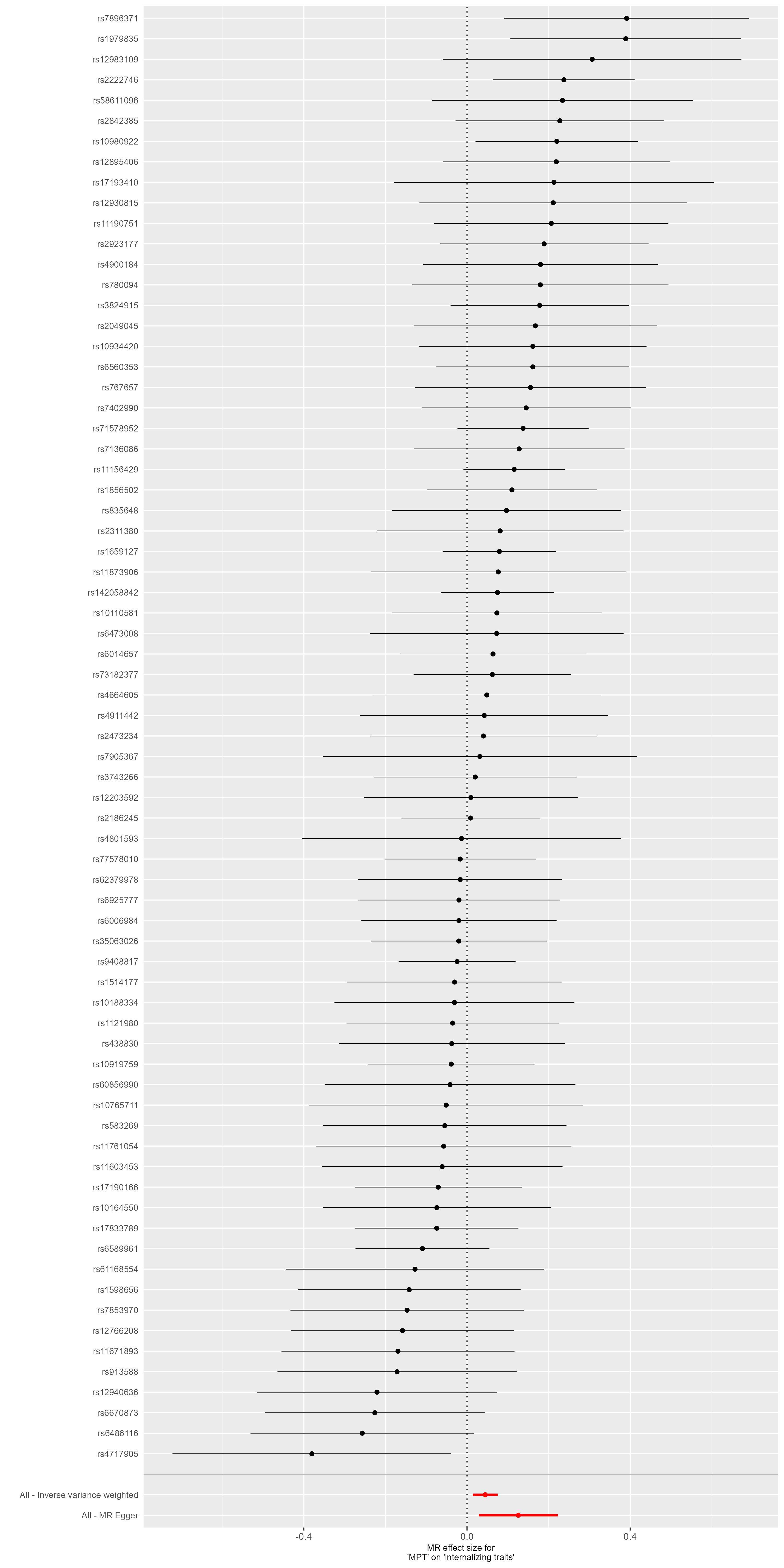

**Supplementary Figure S11 – Forest Plot - Early Life Internalizing Traits.** This forest plot illustrates the results of single-SNP MR analyses with the MR effect estimates for each SNP on the outcome (Internalizing Traits in children and adolescents).

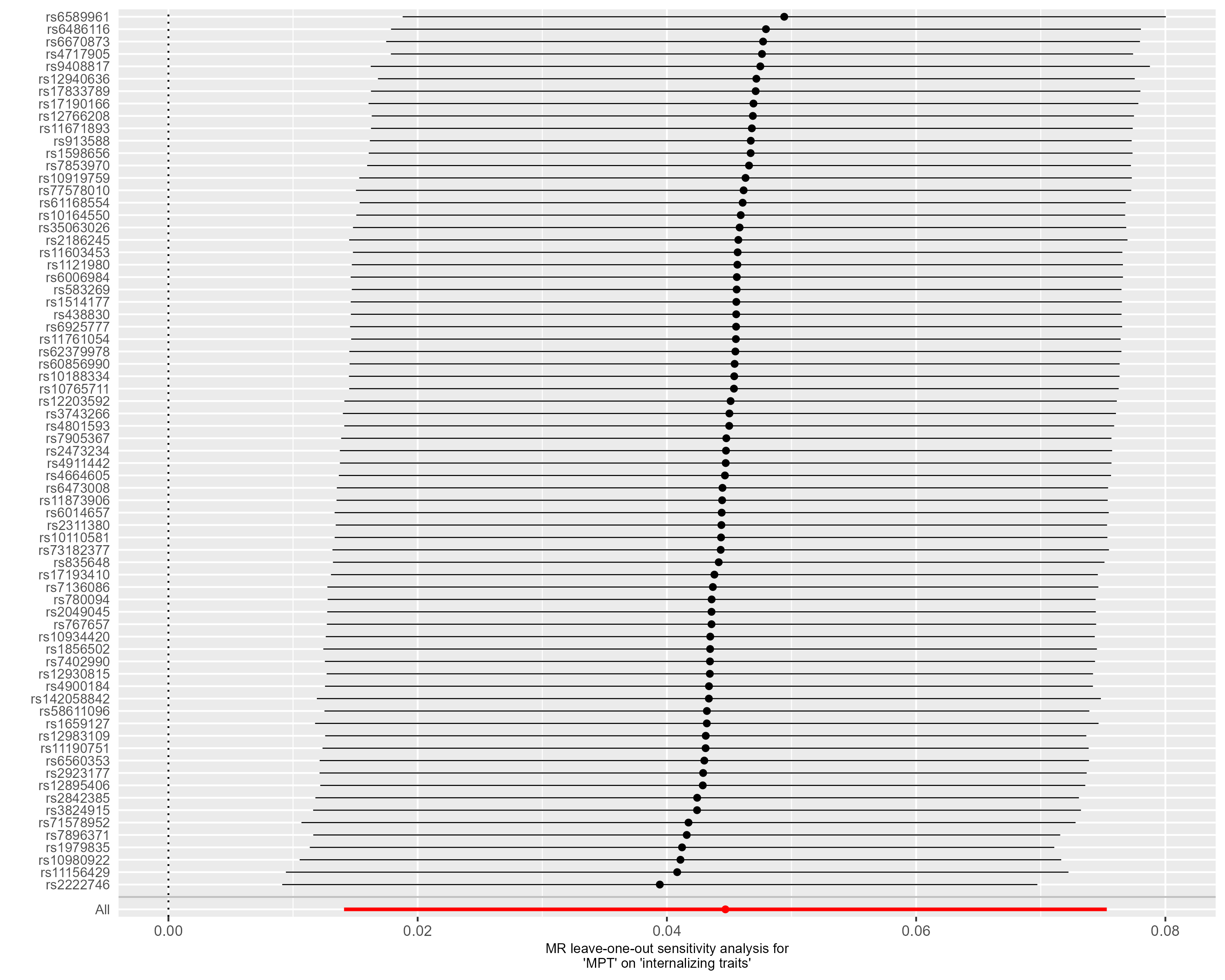

**Supplementary Figure S12 – Leave-one-out analysis - Early Life Internalizing Traits.** This forest plot illustrates the results of MR analyses (IVW) following the exclusion of individual genetic variants from the instrumental variable through leave-one-out analysis.

**Outcome: Depressed Affect**

72 of the 76 target SNPs of male puberty timing were available in the outcome GWAS by Nagel, et al. [18]. One outlier (rs12983109) was identified by MR-PRESSO. Thus, the instrumental variable consisted of 71 genetic variants. The exclusion of the outlier by MR-PRESSO did not alter the results of the primary endpoint (see ‘MR-PRESSO raw’ IVW estimate in Figure 3D). The funnel plot revealed a particularly precise genetic variant (rs11156429) whose effect estimate differed from those obtained by the IVW and MR Egger methods, suggesting a bias due to pleiotropy (see Supplementary Figure S14). Nonetheless, even when this variant was excluded in a leave-one-out analysis, the effect of male puberty timing on the outcome remained significant (refer to Supplementary Figure S16).

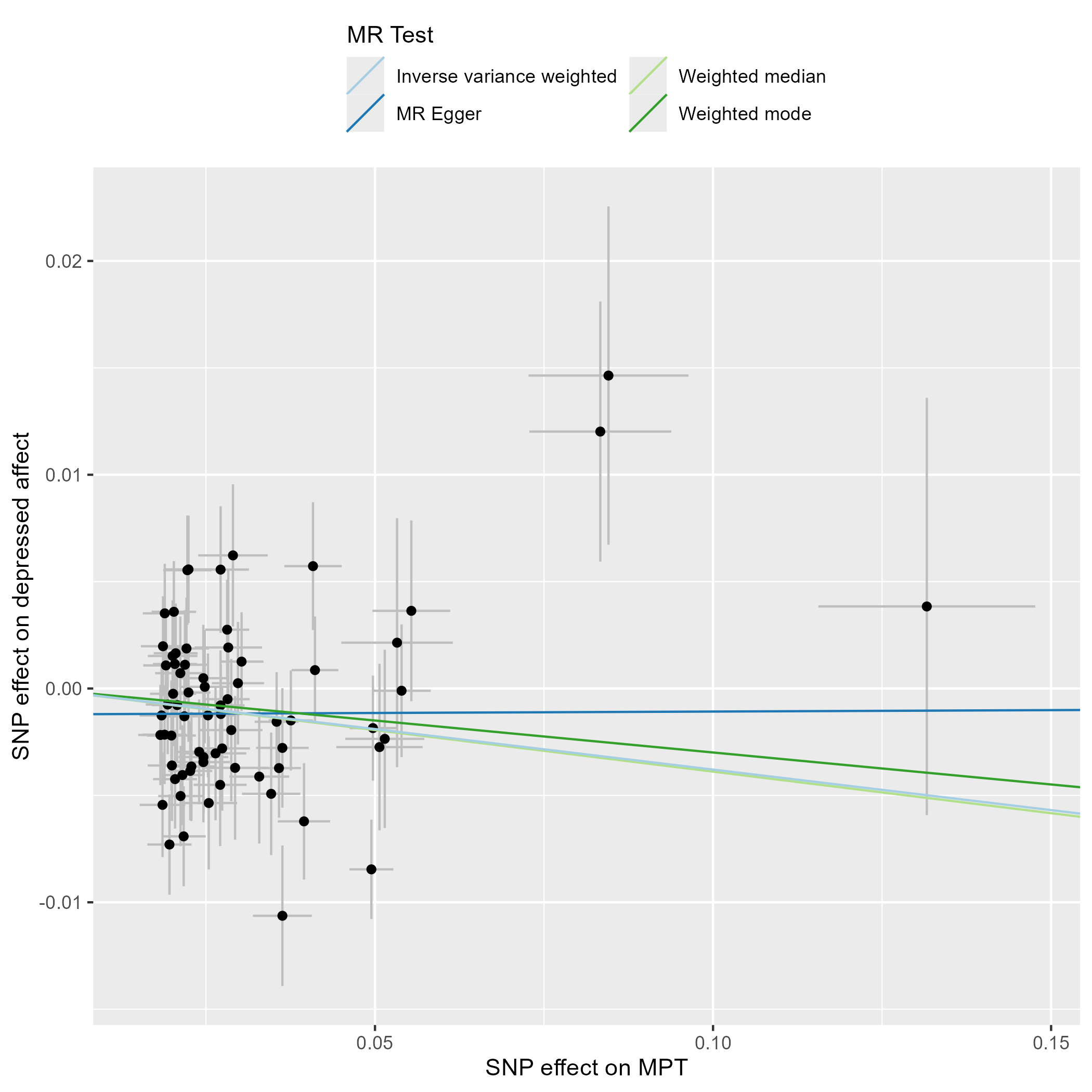

**Supplementary Figure S13 – Scatter Plot – Depressed Affect.** This scatter plot depicts the effect estimates (beta, error bars represent the standard error) of each genetic variant included in the analysis on the exposure (male puberty timing [29]) and the outcome (‘Depressed Affect’ subdomain of Neuroticism, [18]).

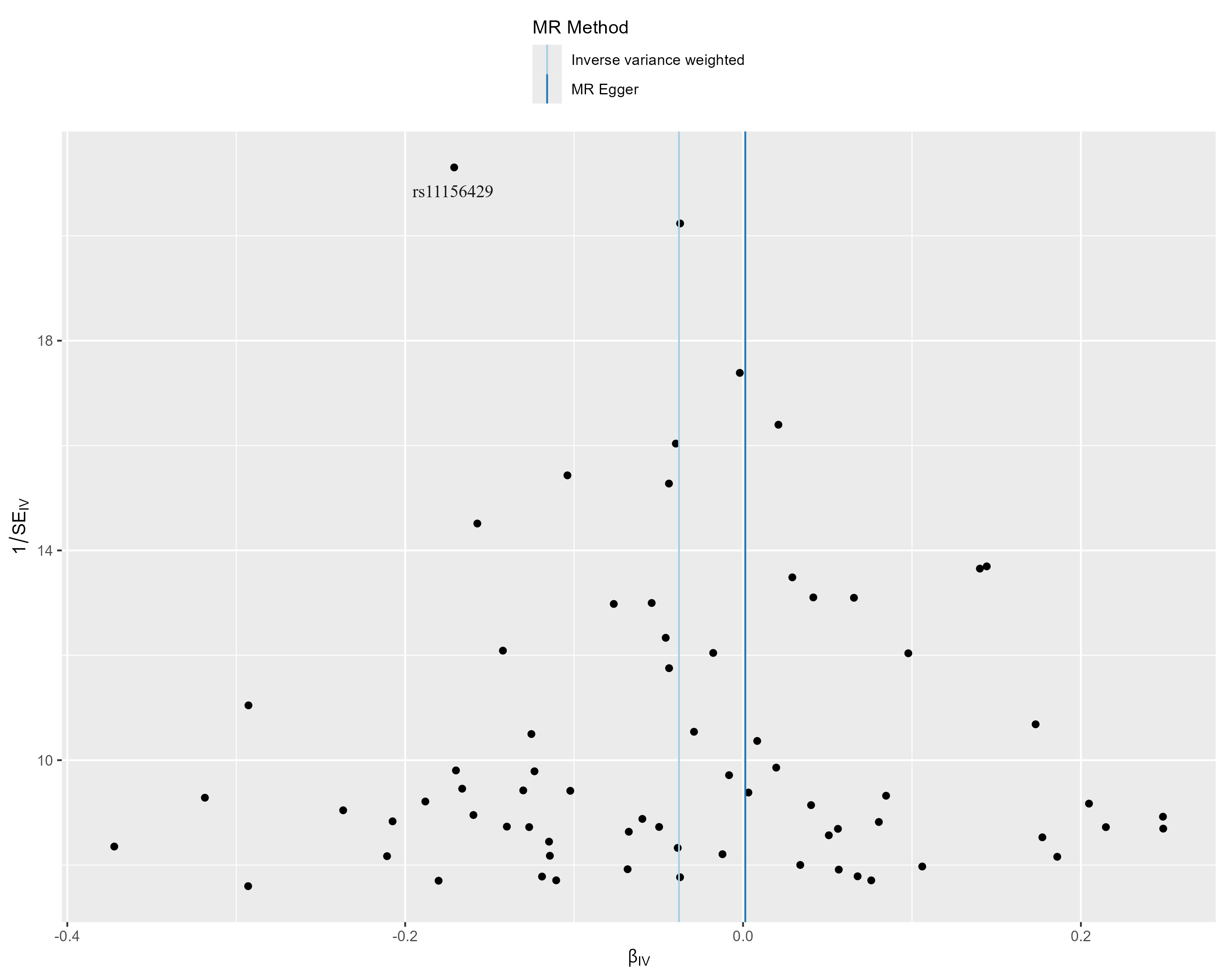

**Supplementary Figure S14 – Funnel Plot - Depressed Affect.** This funnel plot illustrates the precision of each genetic variant (as measured by the inverse of the standard error (SE_IV_)) and their MR effect estimate (β_IV_).

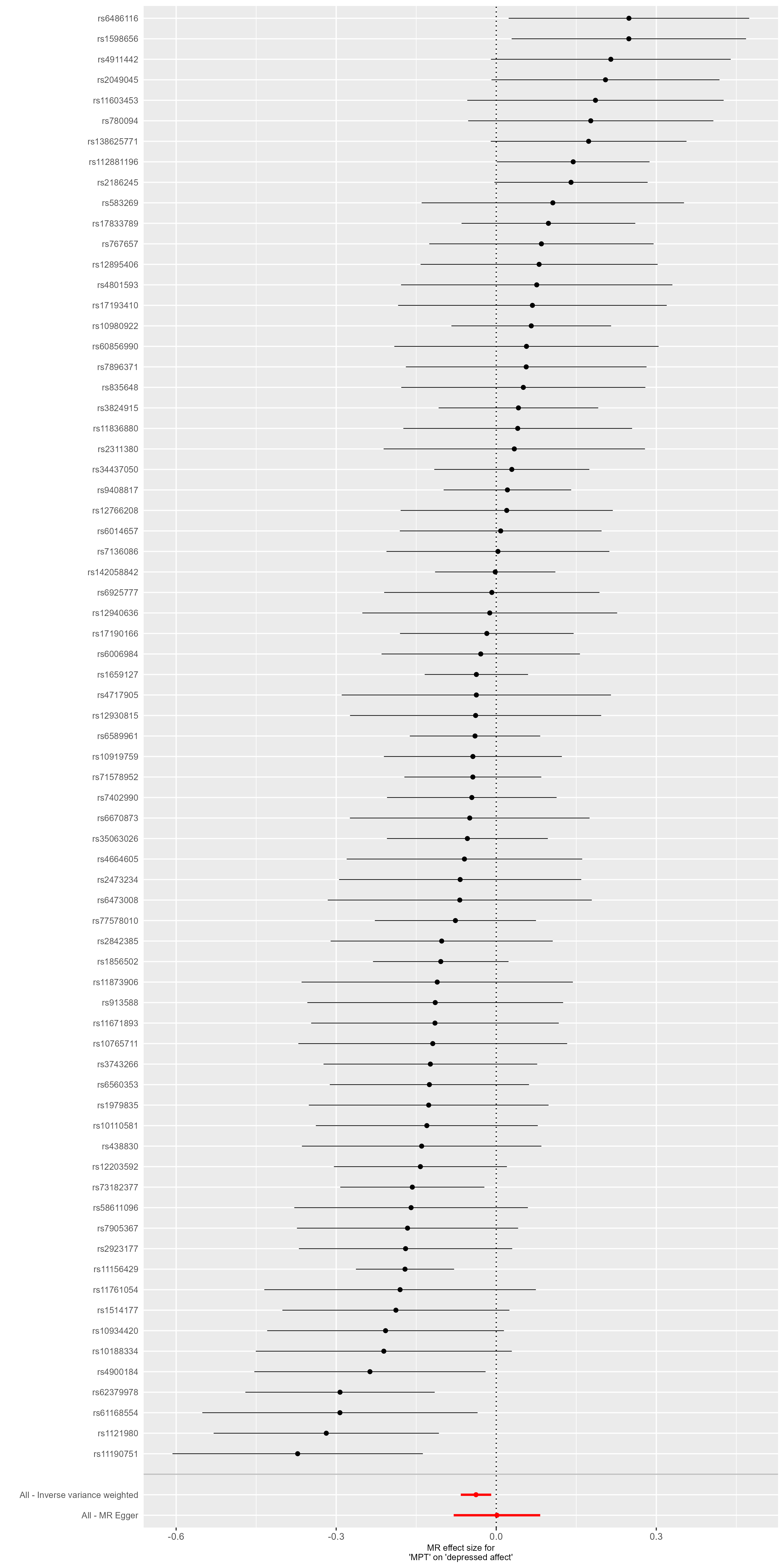

**Supplementary Figure S15 – Forest Plot - Depressed Affect.** This forest plot illustrates the results of single-SNP MR analyses with the MR effect estimates for each SNP on the outcome (‘Depressed Affect’ subdomain of Neuroticism).

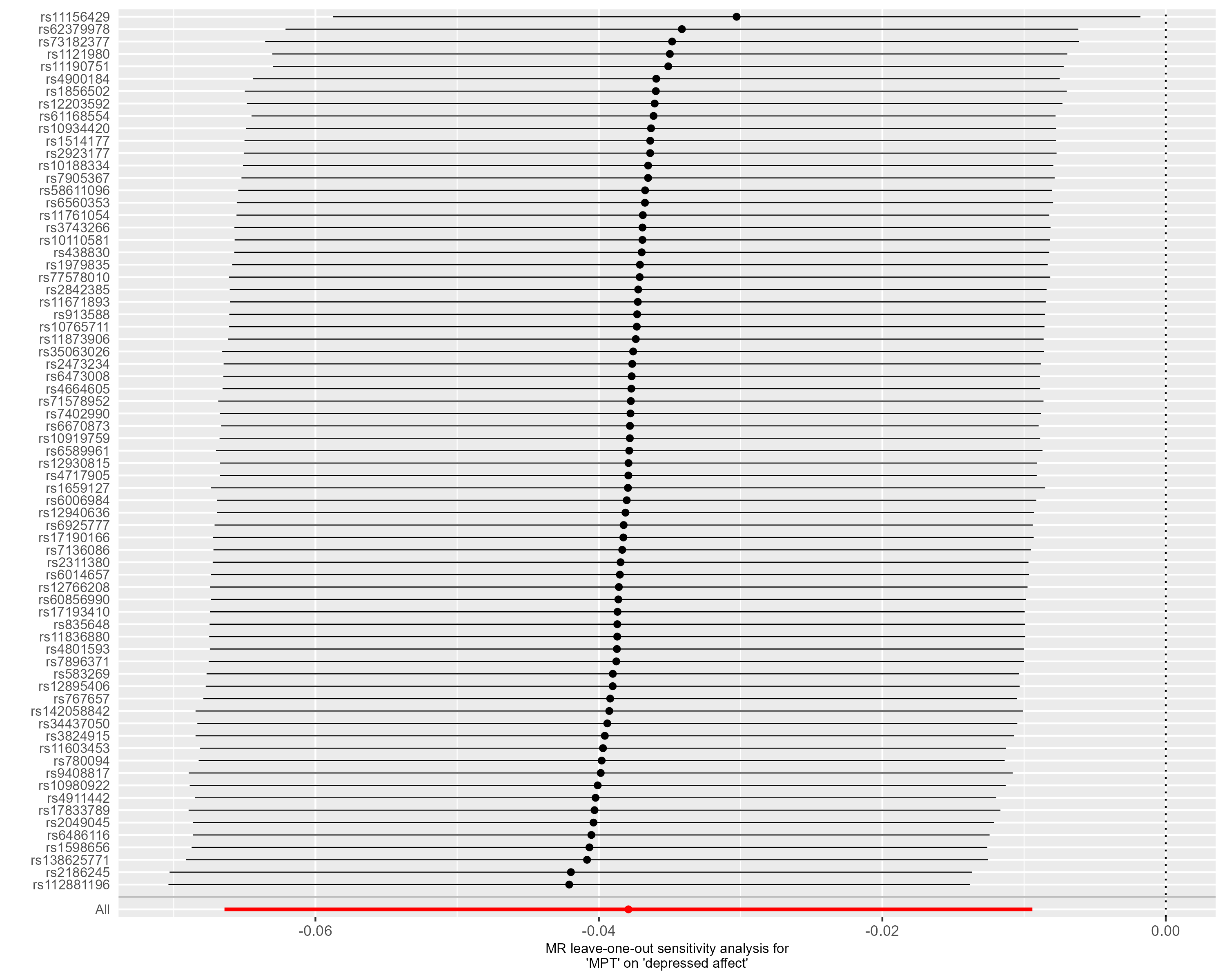

**Supplementary Figure S16 – Leave-one-out analysis - Depressed Affect.** This forest plot illustrates the results of MR analyses (IVW) following the exclusion of individual genetic variants from the instrumental variable through leave-one-out analysis.

**Outcome: Age at Onset of Depression.**

74 of the 76 target SNPs of male puberty timing were available in the outcome GWAS by Nagel, et al. [18]. No outlier was identified by MR-PRESSO. The uncorrected effect (p = 0.034) indicates that a younger onset of male puberty leads to an earlier onset of depression. However, this effect was not maintained after FDR correction (FDR corrected p = 0.115). The Q-statistic did not reveal significant heterogeneity (IVW: df=73, Q-statistic: 91.40, p=0.071; MR Egger: df=72, Q-statistic: 87.54, p=0.103). However, robust methods showed a mixed picture with non-significant estimates with the weighted median, the weighted mode approach, and the BMI-correct IVW estimate (Supplemental Figure S17). In line with this observation, leave-one-out-analyses revealed that after the exclusion of single variants, the 95%-CI of the effect estimate included the null effect (Supplementary Figure S21).

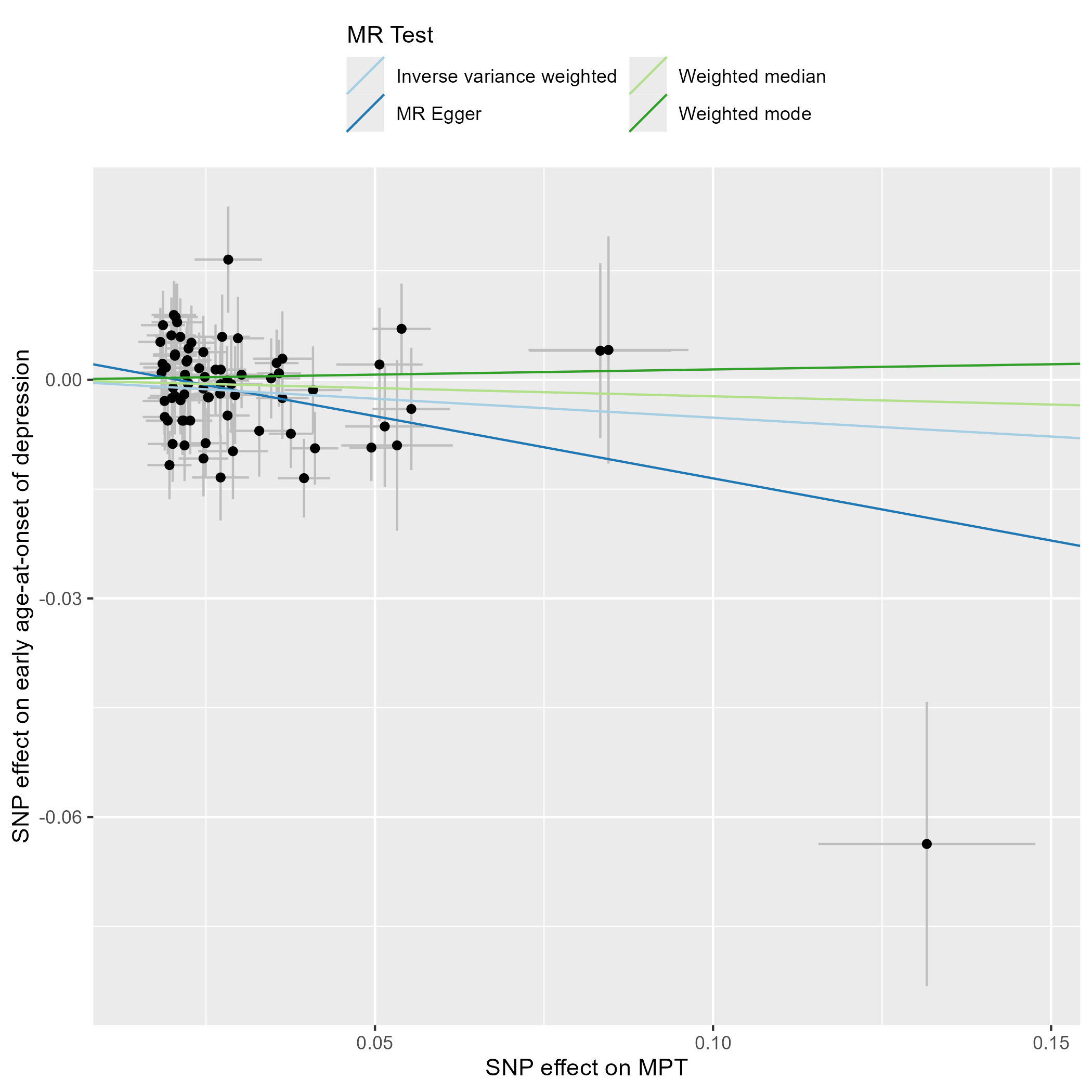

**Supplementary Figure S17 – Scatter Plot – Age at Onset of Depression.** This scatter plot depicts the effect estimates (beta, error bars represent the standard error) of each genetic variant included in the analysis on the exposure (male puberty timing [29]) and the outcome (Age at Onset of Depression [22]).

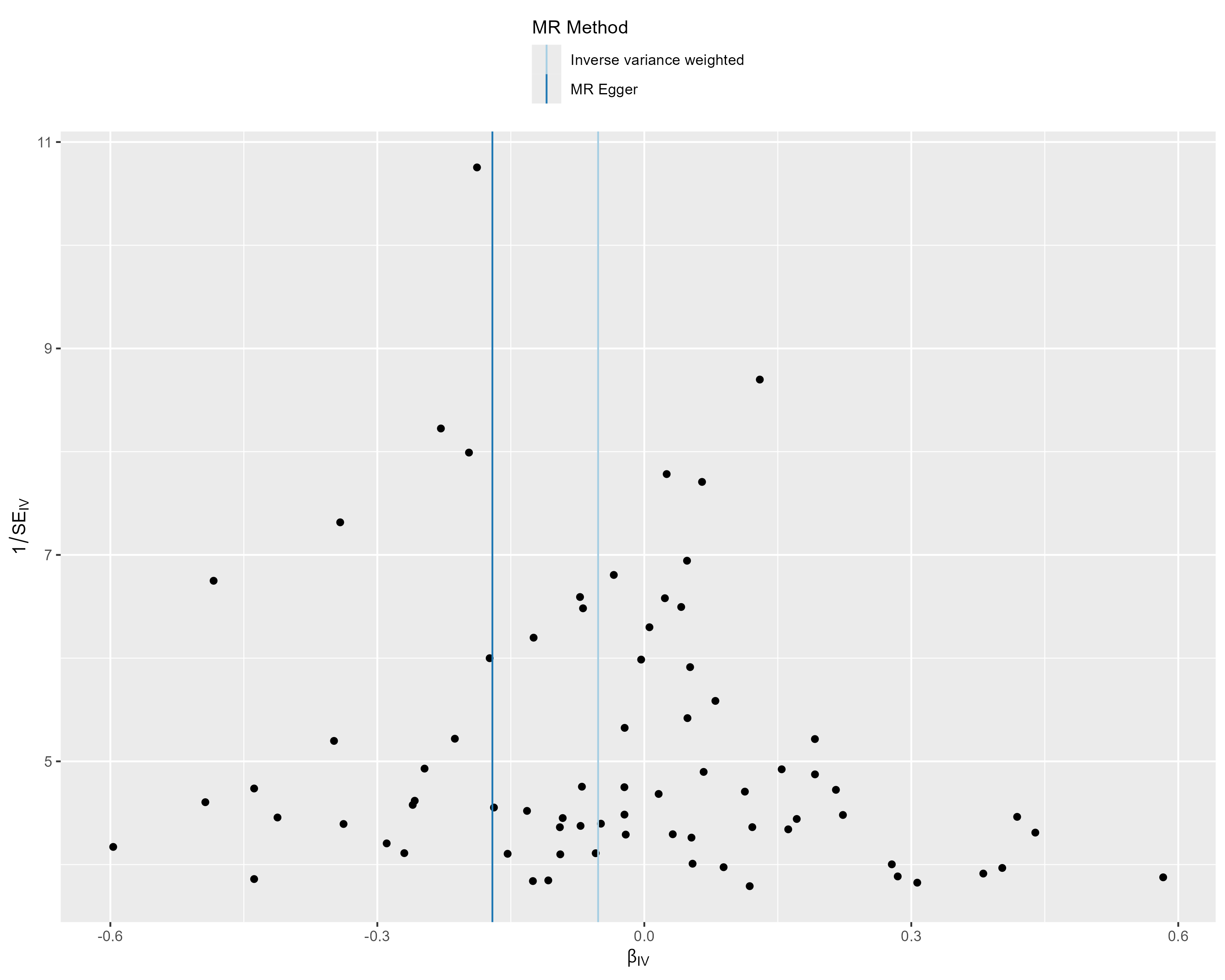

**Supplementary Figure S18 – Funnel Plot - Age at Onset of Depression.** This funnel plot illustrates the precision of each genetic variant (as measured by the inverse of the standard error (SE_IV_)) and their MR effect estimate (β_IV_).

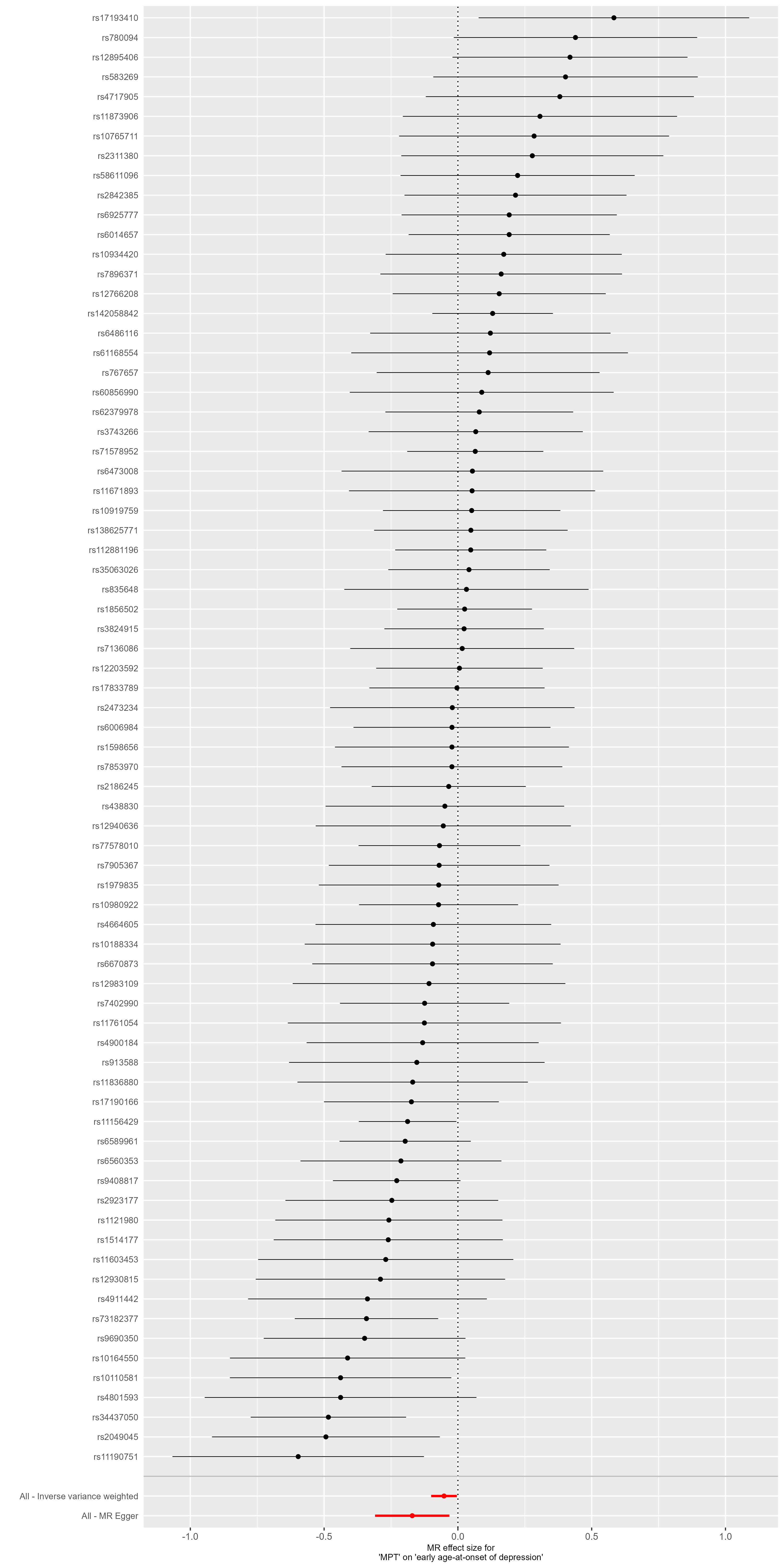

**Supplementary Figure S19 – Forest Plot - Age at Onset of Depression.** This forest plot illustrates the results of single-SNP MR analyses with the MR effect estimates for each SNP on the outcome (Age at Onset of Depression).

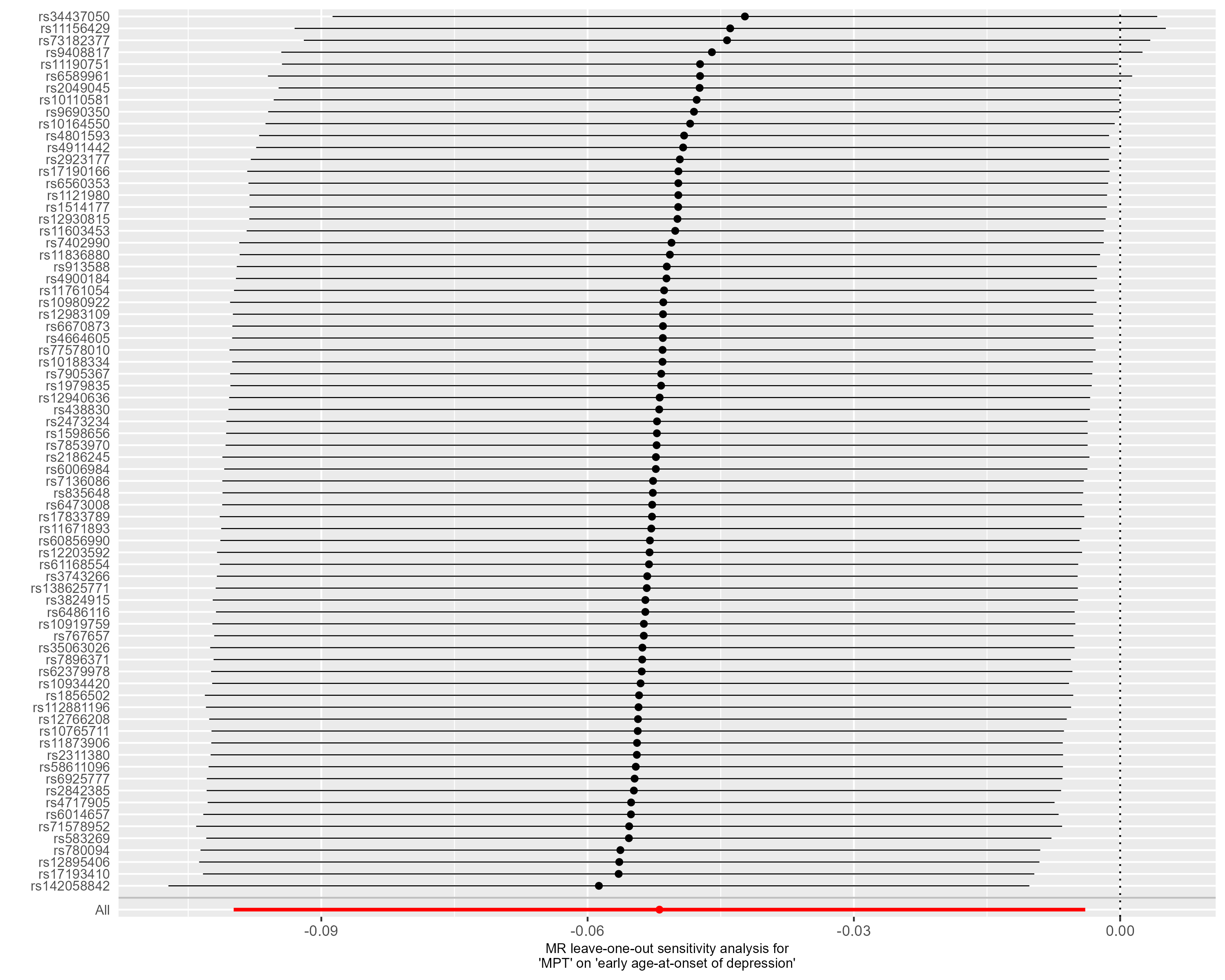

**Supplementary Figure S20 – Leave-one-out analysis - Age at Onset of Depression.** This forest plot illustrates the results of MR analyses (IVW) following the exclusion of individual genetic variants from the instrumental variable through leave-one-out analysis.

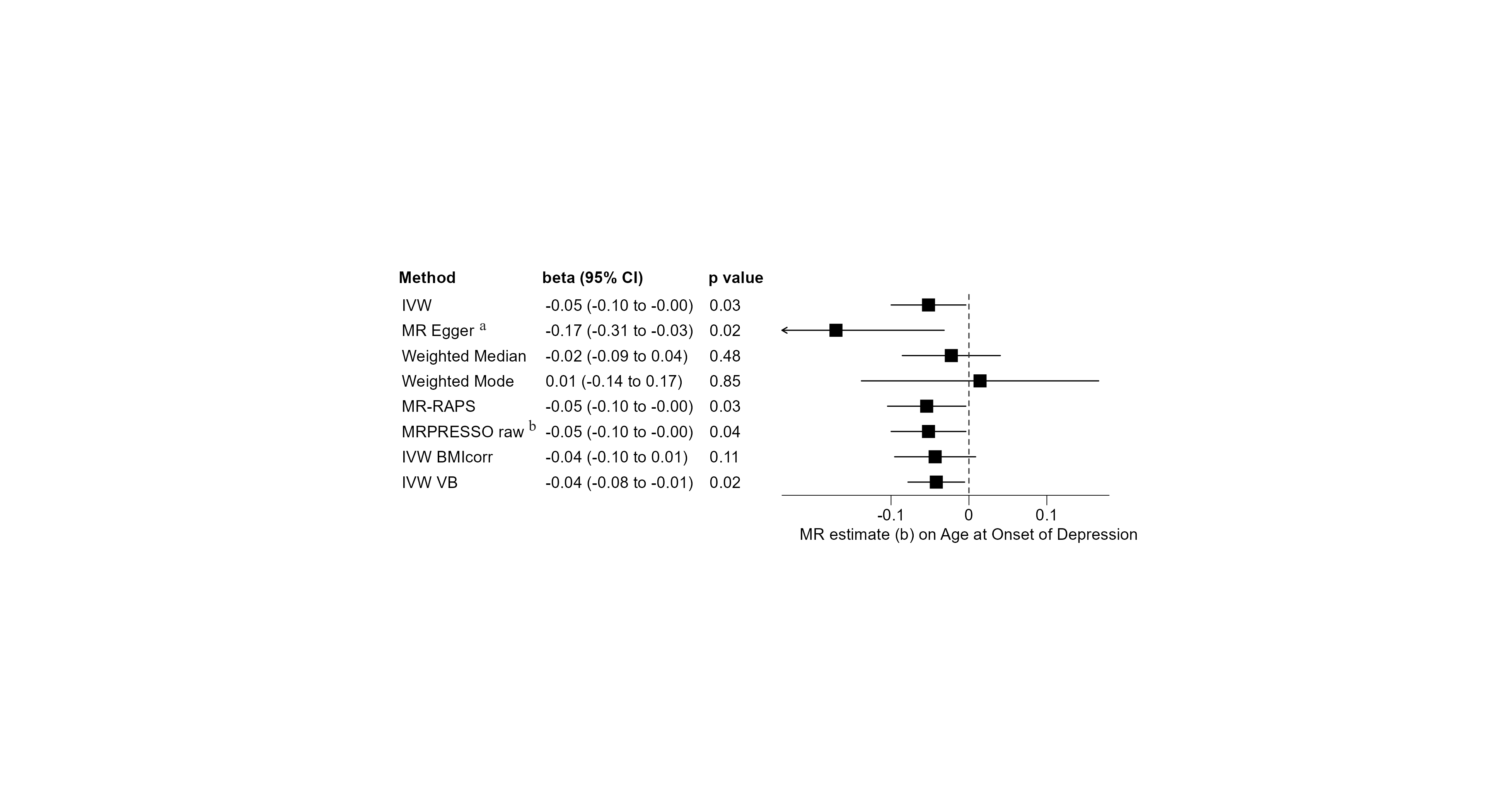

**Supplementary Figure S21 – Sensitivity Analyses – Age at Onset of Depression.** Univariable MR analyses on the effect of male puberty timing on the age at onset of depression. Negative MR estimates indicate that earlier male pubertal timing is associated with younger age at onset of depression. a: Eggers-intercept did not indicate significant directional pleiotropy (intercept=0.0035, se=0.0020, p=0.079). However, the I^2^-statistic was 0.734 and thus well below the threshold of 0.9. This suggests a violation of the NOME assumption, essential to the validity of MR Egger results. Therefore, the MR Egger result should be interpreted with caution. b: The MR-PRESSO global test for pleiotropy was not significant (RSSobs=94.12, p=0.084), and no outliers were identified by MR-PRESSO. Thus, the MR-PRESSO corrected IVW estimate (IVW) corresponds to the uncorrected IVW estimate (MR-PRESSO raw).

**Supplementary Results 1 – Details on outcomes with primary endpoint p value < 0.05**

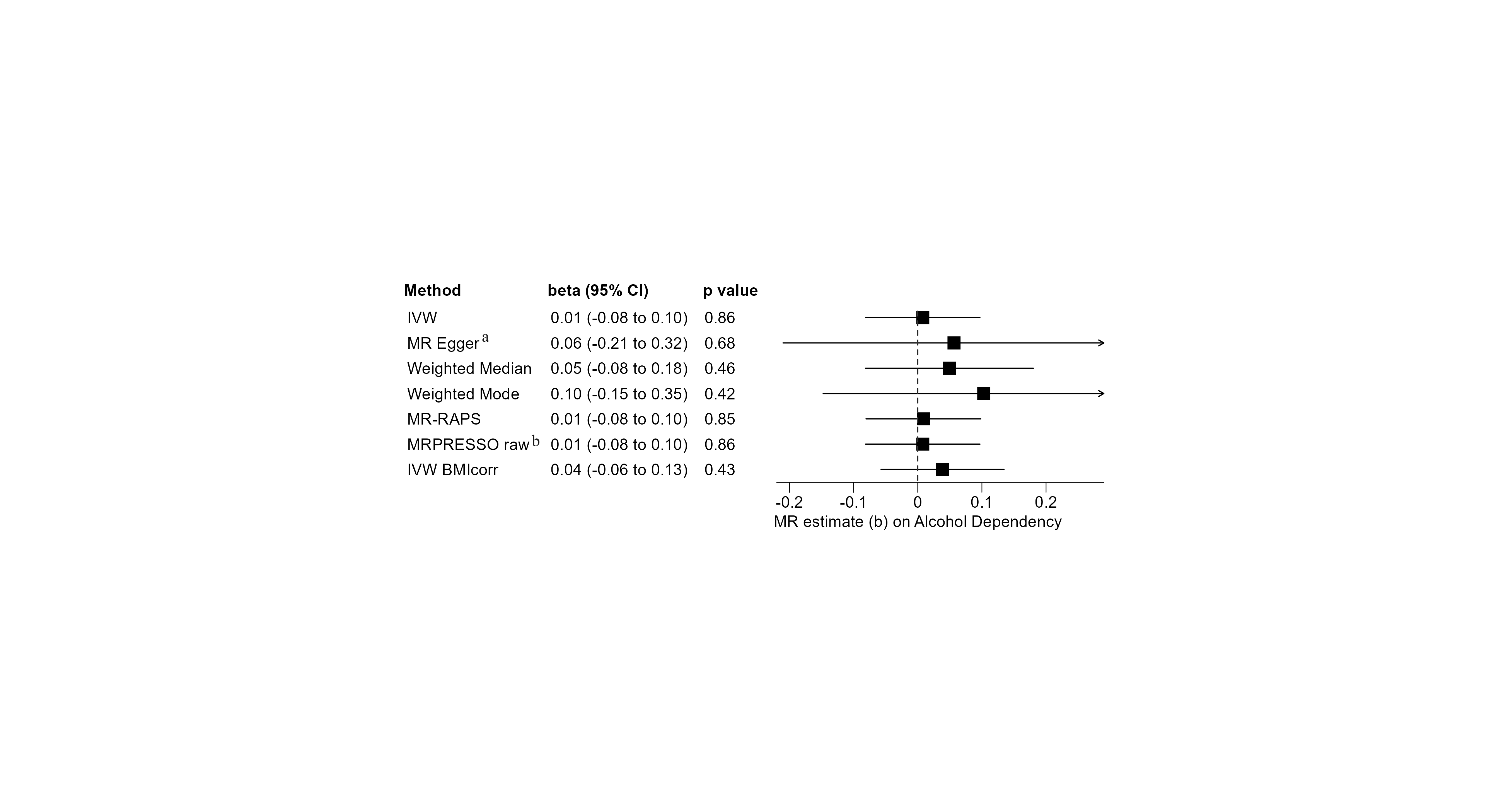

**Supplementary Figure S22 – Sensitivity Analyses – Alcohol Dependency.** Univariable MR analyses on the effect of male puberty timing on the risk of developing alcohol dependence. Seventy-five of the target SNPs for male puberty timing were covered in the outcome GWAS by Walters, et al. [16]. Positive MR estimates indicate that later male puberty timing is associated with higher risk of developing alcohol dependence. The Q-statistic did not reveal significant heterogeneity (IVW: df=74, Q-statistic: 82.60, p=0.231; MR Egger: df=73, Q-statistic: 82.43, p=0.211). a: Eggers-intercept did not indicate significant directional pleiotropy (intercept= -0.0015, se= 0.0038, p=0.705). The I^2^-statistic was 0.787, indicating a violation of the NOME assumption. b: The MR-PRESSO global test for pleiotropy was not significant (RSSobs=84.70, p=0.228), and no outliers were identified by the MR-PRESSO. Thus, the MR-PRESSO corrected IVW estimate (IVW) corresponds to the uncorrected IVW estimate (MR-PRESSO raw).

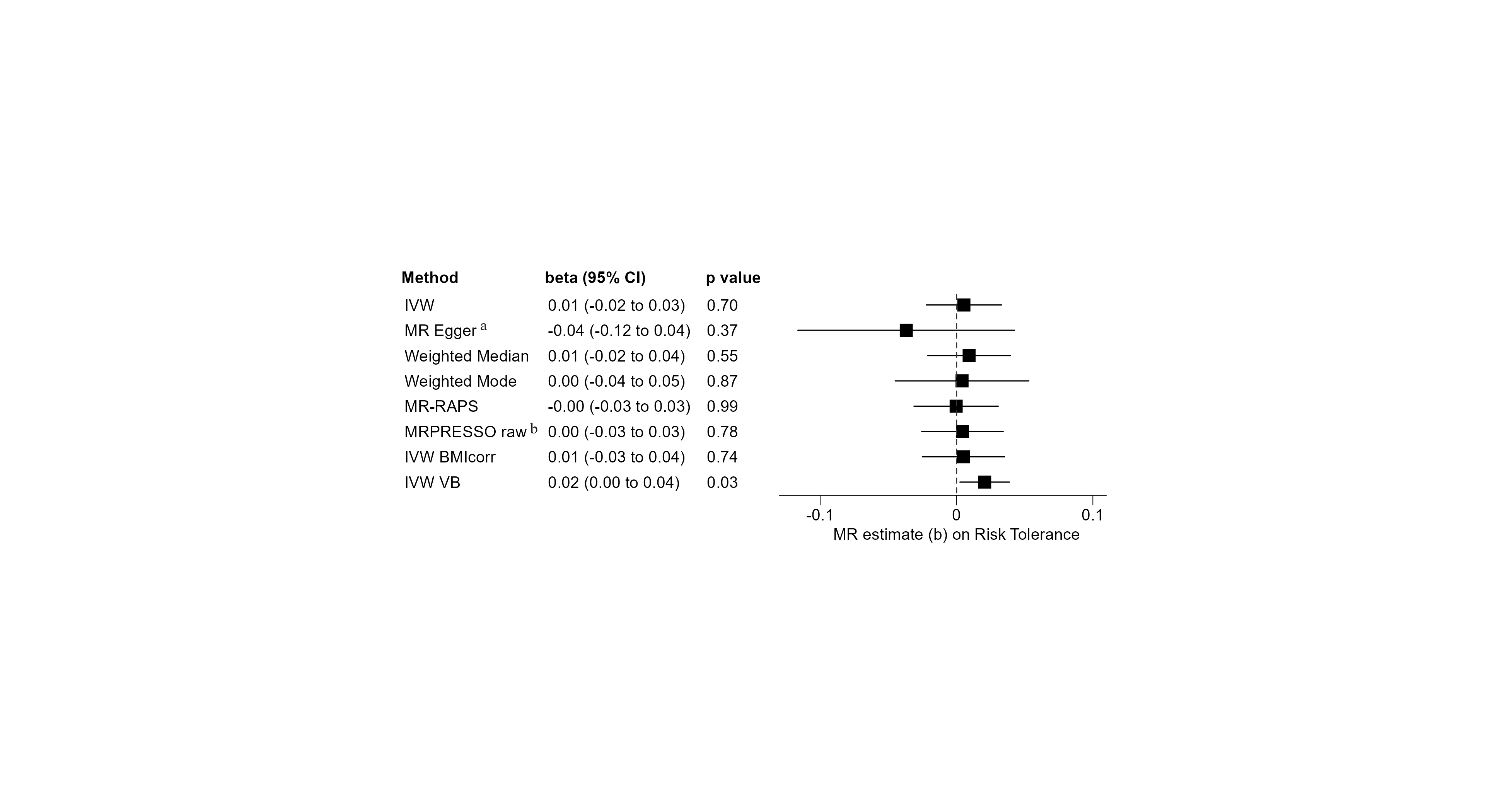

**Supplementary Figure S23 – Sensitivity Analyses – Risk Tolerance.** Univariable MR analyses on the effect of male puberty timing on general risk tolerance. 75 of the 76 target SNPs for male puberty timing were covered in the outcome GWAS by Karlsson Linnér, et al. [13]. Positive MR estimates indicate that later male puberty timing is associated with greater tolerance towards risky behavior. The Q-statistic revealed significant heterogeneity (IVW: df=72, Q-statistic: 147.20, p= 4x10^-7^; MR Egger: df=71, Q-statistic: 144.68, p=5x10^-7^). a: Eggers-intercept did not indicate significant directional pleiotropy (intercept= 0.0013, se= 0.0012, p=0.270). The I^2^-statistic was 0.747, and thus well below the threshold of 0.9, indicating a violation of the NOME assumption. Therefore, the MR Egger result should be interpreted with caution. b: The MR-PRESSO global test for pleiotropy was significant (RSSobs=184.55, p<3x10^-4^), and two outliers were identified by MR-PRESSO (see Supplemental Table S2 for details). Thus, the instrumental variable consisted of 73 SNPs. As the effect estimate based on the exposure ‘Voice Break’ indicated an effect on ‘Risk Tolerance’, additional sensitivity analyses were undertaken with ‘Voice Break’ as an exposure (not shown in the figure), all with non-significant findings: MR-PRESSO raw (beta=0.02, 95%-CI [-0.01, 0.04], p=0.188), MR Egger (beta=-0.01, 95%-CI [-0.04, 0.06], p=0.677), Weighted Median (beta=0.01, 95%-CI [-0.01, 0.03], p=0.450), and Weighted Mode (beta=0.00, 95%-CI [-0.03, 0.04], p=0.829).

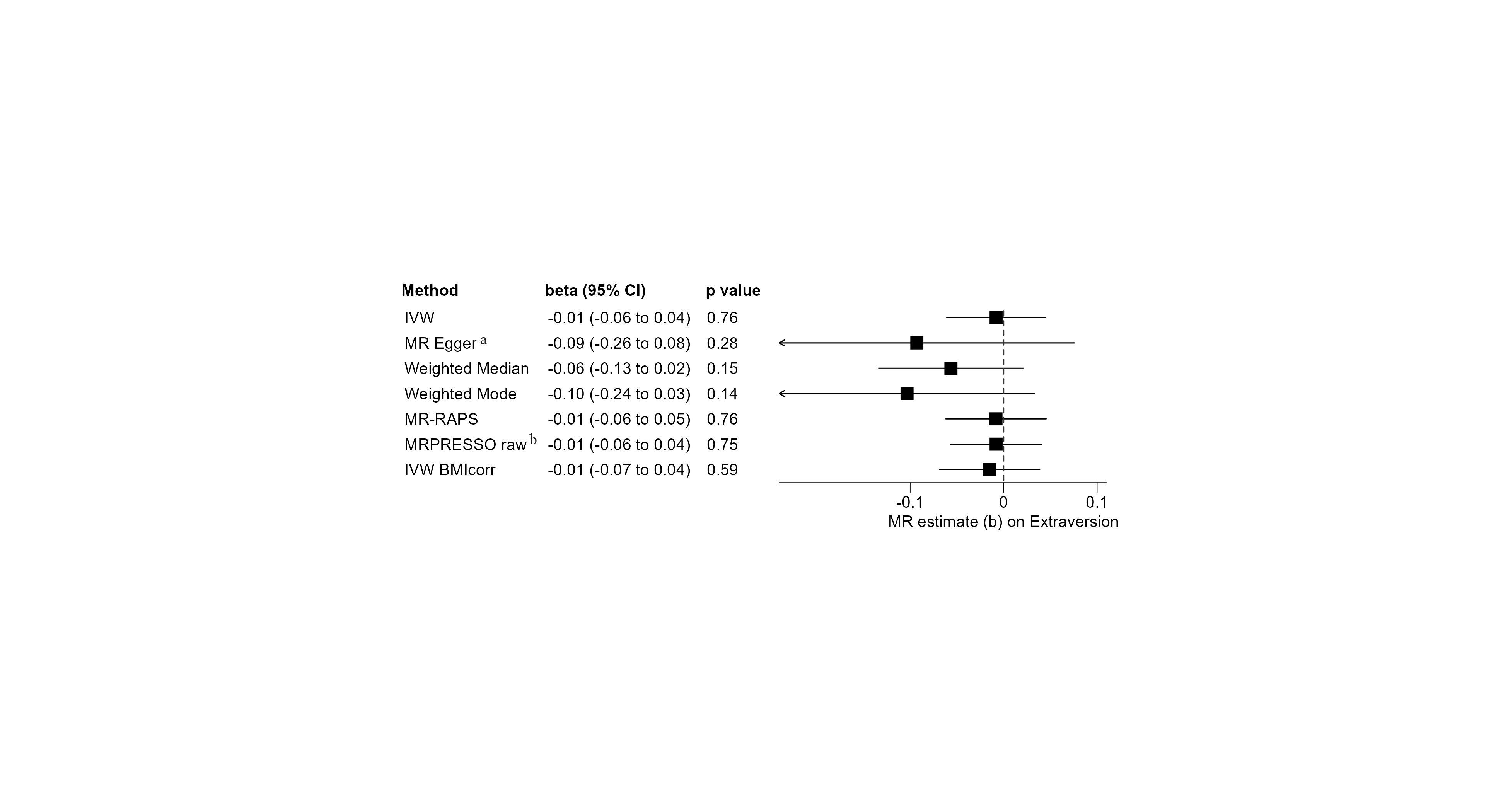

**Supplementary Figure S24 – Sensitivity Analyses – Extraversion.** Univariable MR analyses on the effect of male puberty timing on extraversion. 73 of the 76 target SNPs for male puberty timing were covered in the outcome GWAS by Van den Berg, et al. [10]. Positive MR estimates indicate that later male puberty timing is associated with greater expression of the character trait ‘extraversion’. The Q-statistic did not reveal significant heterogeneity (IVW: df=72, Q-statistic: 62.12, p=0.790; MR Egger: df=71, Q-statistic: 61.05, p=0.794). a: Eggers-intercept did not indicate significant directional pleiotropy (intercept= 0.0025, se=0.0024, p=0.303). The I^2^-statistic was 0.917, indicating no major violation of the NOME assumption for the conduction of MR Egger. b: The MR-PRESSO global test for pleiotropy was not significant (RSSobs=63.92, p=0.790), and no outliers were identified by MR-PRESSO (see Supplemental Table S2 for details). Thus, the MR-PRESSO corrected IVW estimate (IVW) corresponds to the uncorrected IVW estimate (MR-PRESSO raw).

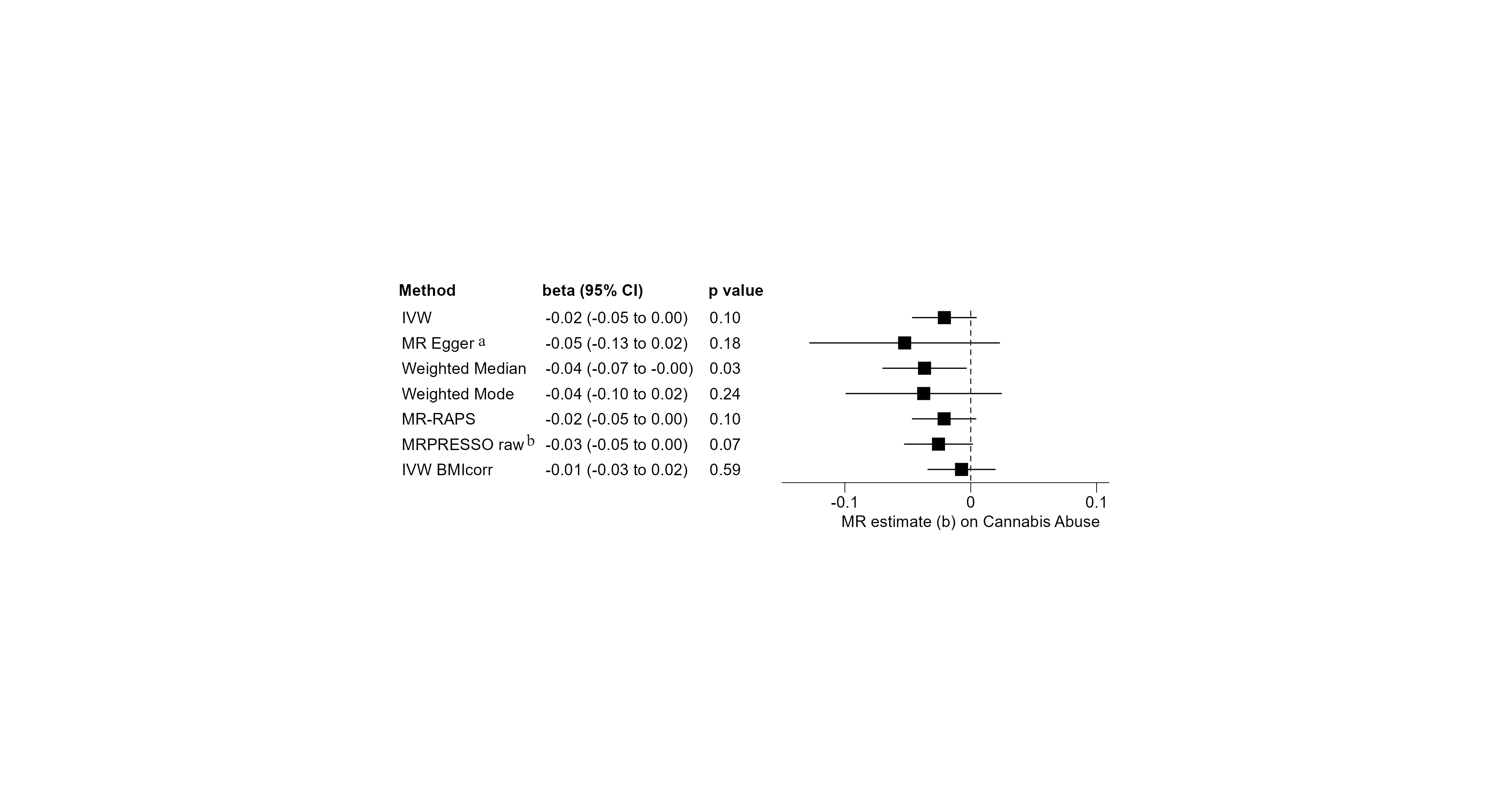

**Supplementary Figure S25 – Sensitivity Analyses – Cannabis Abuse.** Univariable MR analyses on the effect of male puberty timing on cannabis abuse. All 76 target SNPs for male puberty timing were covered in the outcome GWAS by Johnson, et al. [15]. Positive MR estimates indicate that later male puberty timing is associated with a higher risk of developing cannabis abuse. The Q-statistic did reveal a small amount of, however significant, heterogeneity (IVW: df=74, Q-statistic: 96.80, p=0.039; MR Egger: df=73, Q-statistic: 95.79, p=0.038). a: Eggers-intercept did not indicate significant directional pleiotropy (intercept=0.0009, se=0.0011, p=0.386). The I^2^-statistic was 0.783, indicating a relevant violation of the NOME assumption for the conduction of MR Egger. b: The MR-PRESSO global test for pleiotropy was significant (RSSobs=115.37, p=0.003), and one outlier was identified by the MR-PRESSO method (see Supplemental Table S2 for details). Thus, the instrumental variable consisted of 75 SNPs.

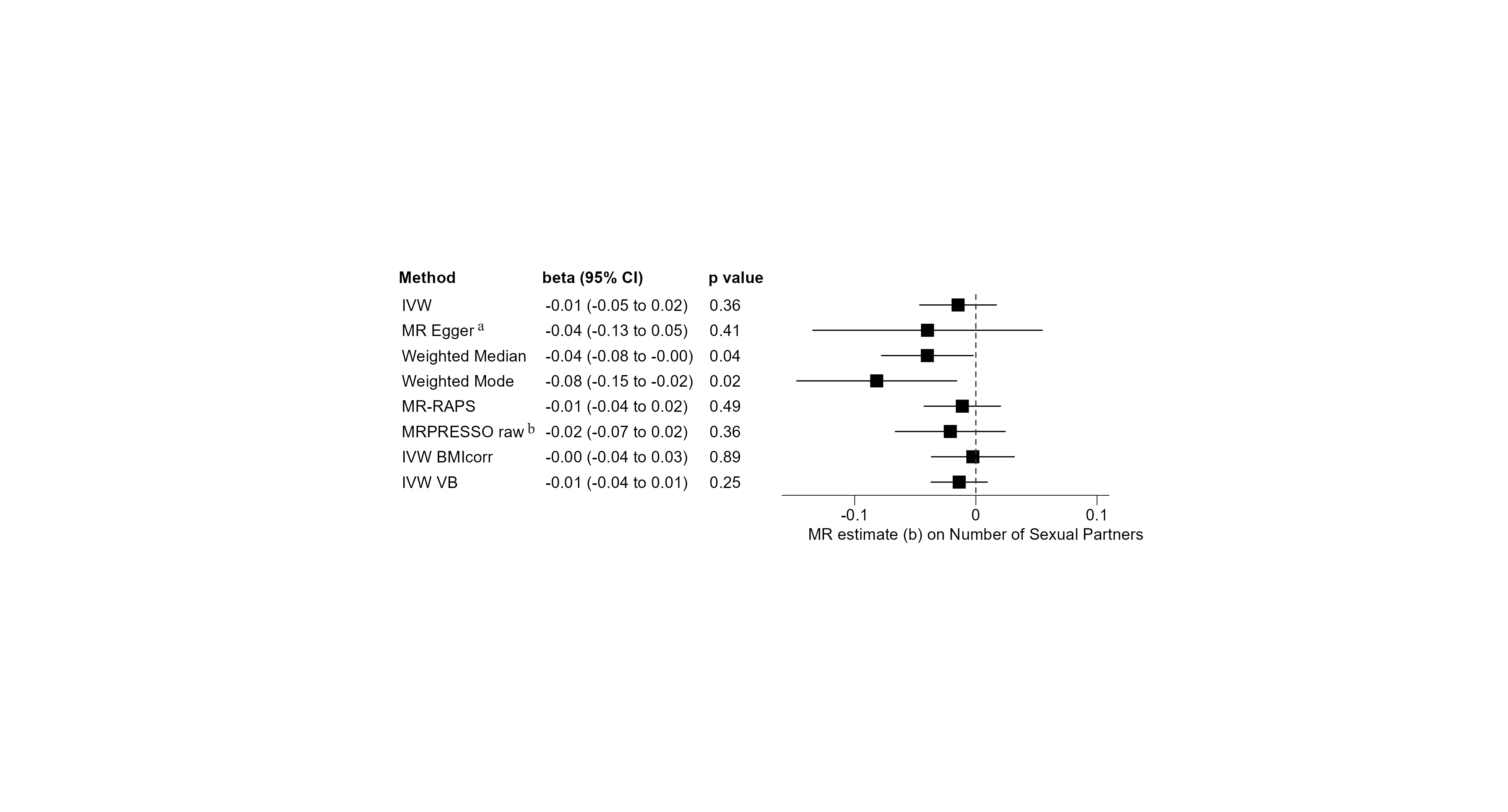

**Supplementary Figure S26 – Sensitivity Analyses – Number of Sexual Partners.** Univariable MR analyses on the effect of male puberty timing on the total number of sexual partners. 75 of the 76 target SNPs for male puberty timing were covered in the outcome GWAS by Karlsson Linnér, et al. [13]. Positive MR estimates indicate that later male puberty timing is associated with a larger number of total sexual partners. The Q-statistic did reveal significant heterogeneity (IVW: df=64, Q-statistic: 117.16, p=5x10^-5^; MR Egger: df=63, Q-statistic: 116.60, p=4x10^-4^). a: Eggers-intercept did not indicate significant directional pleiotropy (intercept=0.0075, se=0.00135, p=0.581). The I^2^-statistic was 0.753, and thus well below the threshold of 0.9, indicating a violation of the NOME assumption. Therefore, the MR Egger result should be interpreted with caution. b: The MR-PRESSO global test for pleiotropy was significant (RSSobs=338.26, p<3x10^-4^) and ten outliers were identified by the MR-PRESSO method (see Supplemental Table S2 for details). Thus, the instrumental variable consisted of 65 SNPs.

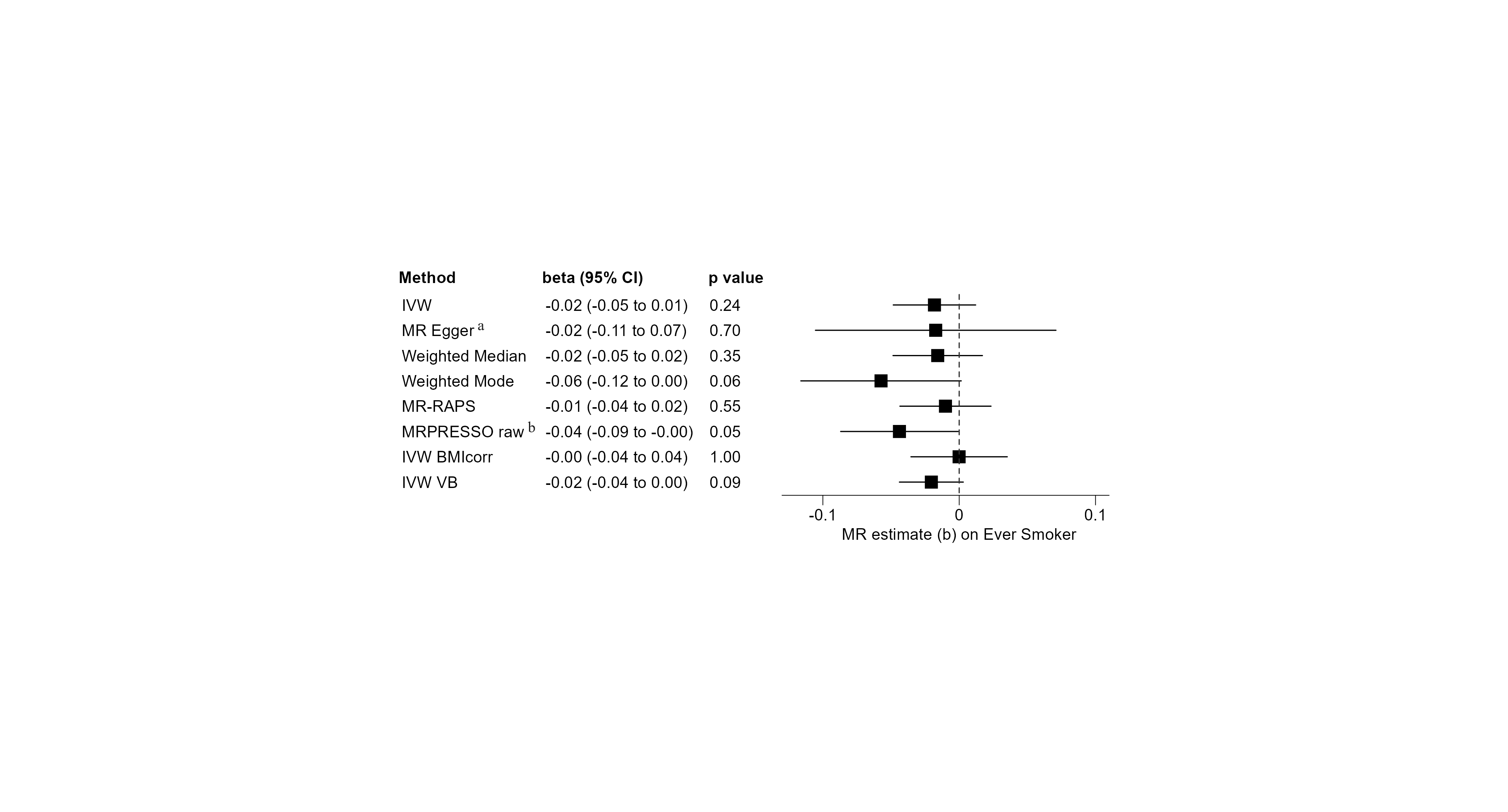

**Supplementary Figure S27 – Sensitivity Analyses – Ever Smoker.** Univariable MR analyses on the effect of male puberty timing on reporting to have ever smoked. 75 of the 76 target SNPs for male puberty timing were covered in the outcome GWAS by Karlsson Linnér, et al. [13]. Positive MR estimates indicate that later male puberty timing is associated with a higher risk of ever initiating smoking. The Q-statistic did reveal significant heterogeneity (IVW: df=66, Q-statistic: 149.32, p=2x10^-8^; MR Egger: df=65, Q-statistic: 149.32, p=1x10^-8^). a: Eggers-intercept did not indicate significant directional pleiotropy (intercept=0.0000, se=0.0013, p=0.983). The I^2^-statistic was 0.744, and thus well below the threshold of 0.9, indicating a violation of the NOME assumption. Therefore, the MR Egger result should be interpreted with caution. b: The MR-PRESSO global test for pleiotropy was significant (RSSobs=386.07, p<3x10^-4^), and eight outliers were identified by MR-PRESSO (see Supplemental Table S2 for details). Thus, the instrumental variable consisted of 67 SNPs. As the IVW estimate without exclusion of pleiotropic variants (MR-PRESSO raw) yielded a (borderline) significant result (p=0.049992), additional sensitivity analyses were undertaken without exclusion of pleiotropic variants, all with non-significant findings (not shown in the figure): MR Egger (beta=-0.03, 95%-CI [-0.16, 0.09], p=0.607), weighted Median (beta=-0.02, 95%-CI [-0.05, 0.01], p=0.275), and Weighted Mode (beta=-0.05, 95%-CI[-0.11, 0.01], p=0.125).

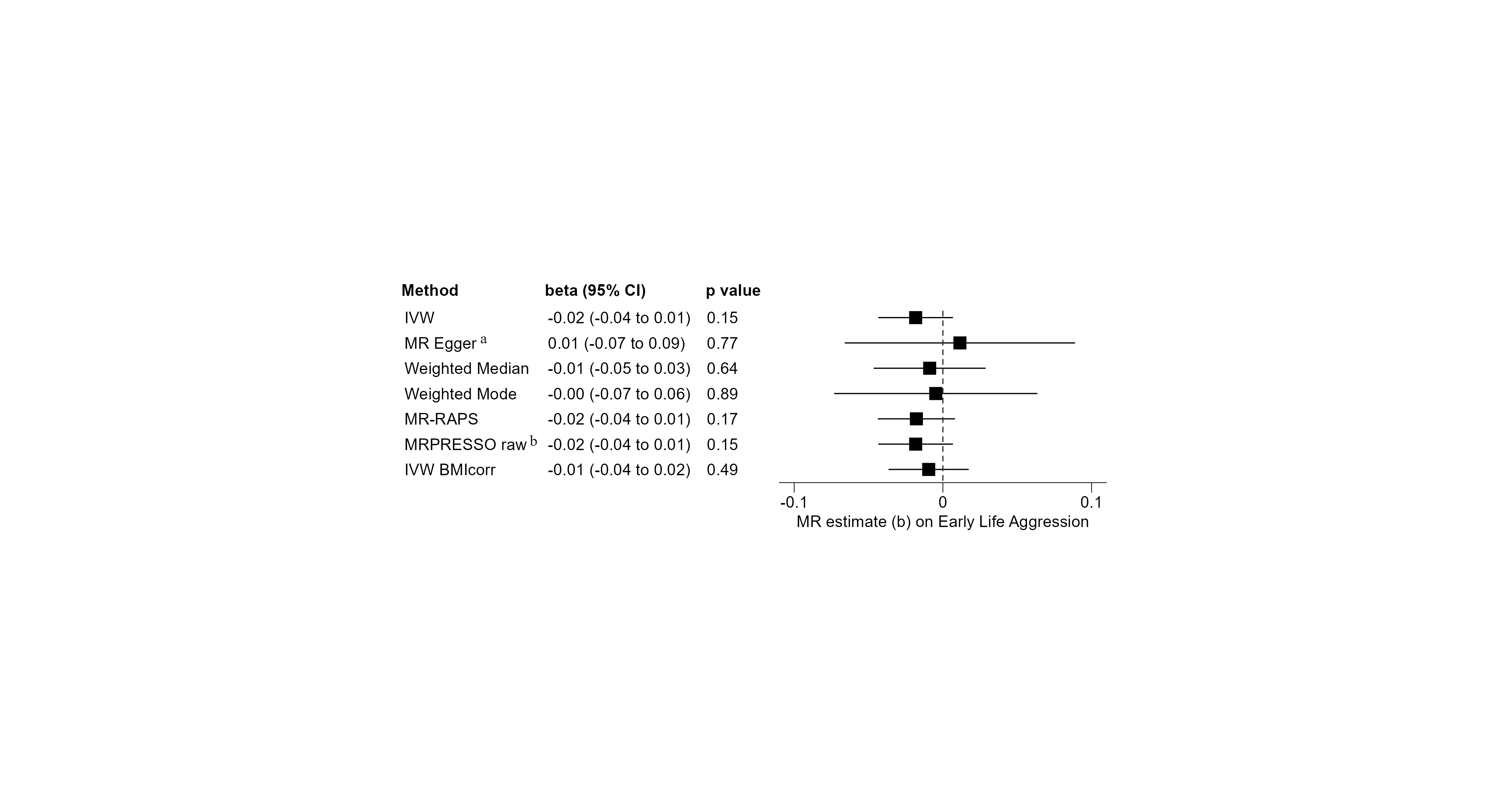

**Supplementary Figure S28 – Sensitivity Analyses – Early Life Aggression.** Univariable MR analyses on the effect of male puberty timing on aggression in children and adolescents. 74 of the 76 target SNPs for male puberty timing were covered in the outcome GWAS by Ip, et al. [12]. Positive MR estimates indicate that later male puberty timing is associated with a higher rating of aggression in childhood and adolescence. The Q-statistic did not reveal significant heterogeneity (IVW: df=73, Q-statistic: 76.98, p=0.352; MR Egger: df=72, Q-statistic: 76.30, p=0.342). a: Eggers-intercept did not indicate significant directional pleiotropy (intercept=-0.0009, se=0.0011, p=0.427). I^2^-statistic was 0.826, and thus below the threshold of 0.9, indicating a violation of the NOME assumption for the conduction of MR Egger. Therefore, the MR Egger result should be interpreted with caution. b: The MR-PRESSO global test for pleiotropy was not significant (RSSobs=79.07, p=0.379), and no outlier was identified by MR-PRESSO (see Supplemental Table S2 for details). Thus, the MR-PRESSO corrected IVW estimate (IVW) corresponds to the uncorrected IVW estimate (MR-PRESSO raw).

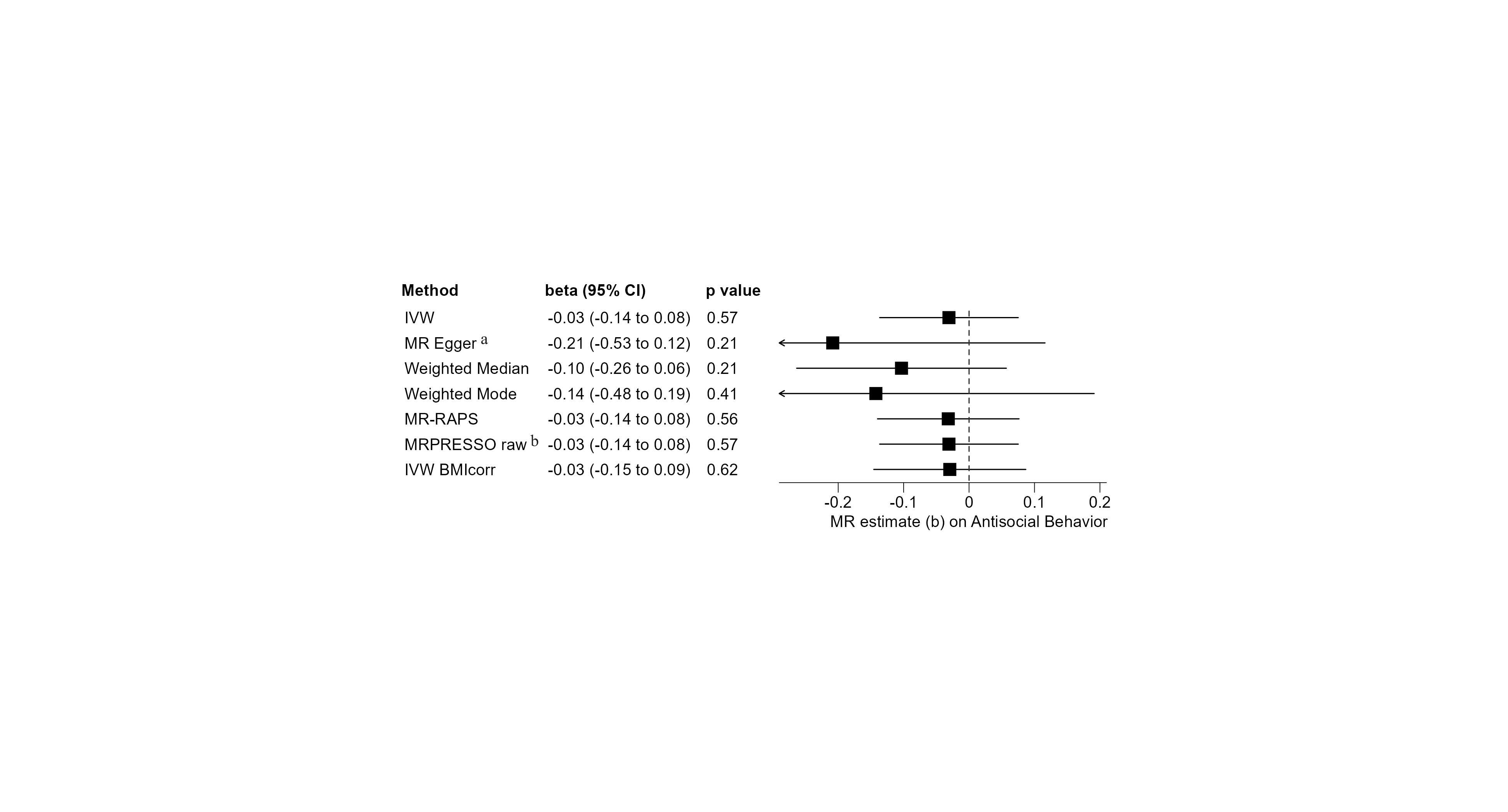

**Supplementary Figure S29 – Sensitivity Analyses – Antisocial Behavior.** Univariable MR analyses on the effect of male puberty timing on antisocial behavior. 75 of the 76 target SNPs for male puberty timing were covered in the outcome GWAS by Tielbeek, et al. [11]. Positive MR estimates indicate that later male puberty timing is associated with more antisocial behavior. The Q-statistic did not reveal significant heterogeneity (IVW: df=74, Q-statistic: 75.70, p=0.423; MR Egger: df=73, Q-statistic: 74.39, p=0.433). a: Eggers-intercept did not indicate significant directional pleiotropy (intercept=0.0053, se=0.0046, p=0.260). I^2^-statistic was 0.822, indicating a relevant violation of the NOME assumption for the conduction of MR Egger. b: The MR-PRESSO global test for pleiotropy was not significant (RSSobs=78.21, p=0.411), and no outlier was identified by MR-PRESSO (see Supplemental Table S2 for details). Thus, the MR-PRESSO corrected IVW estimate (IVW) corresponds to the uncorrected IVW estimate (MR-PRESSO raw).

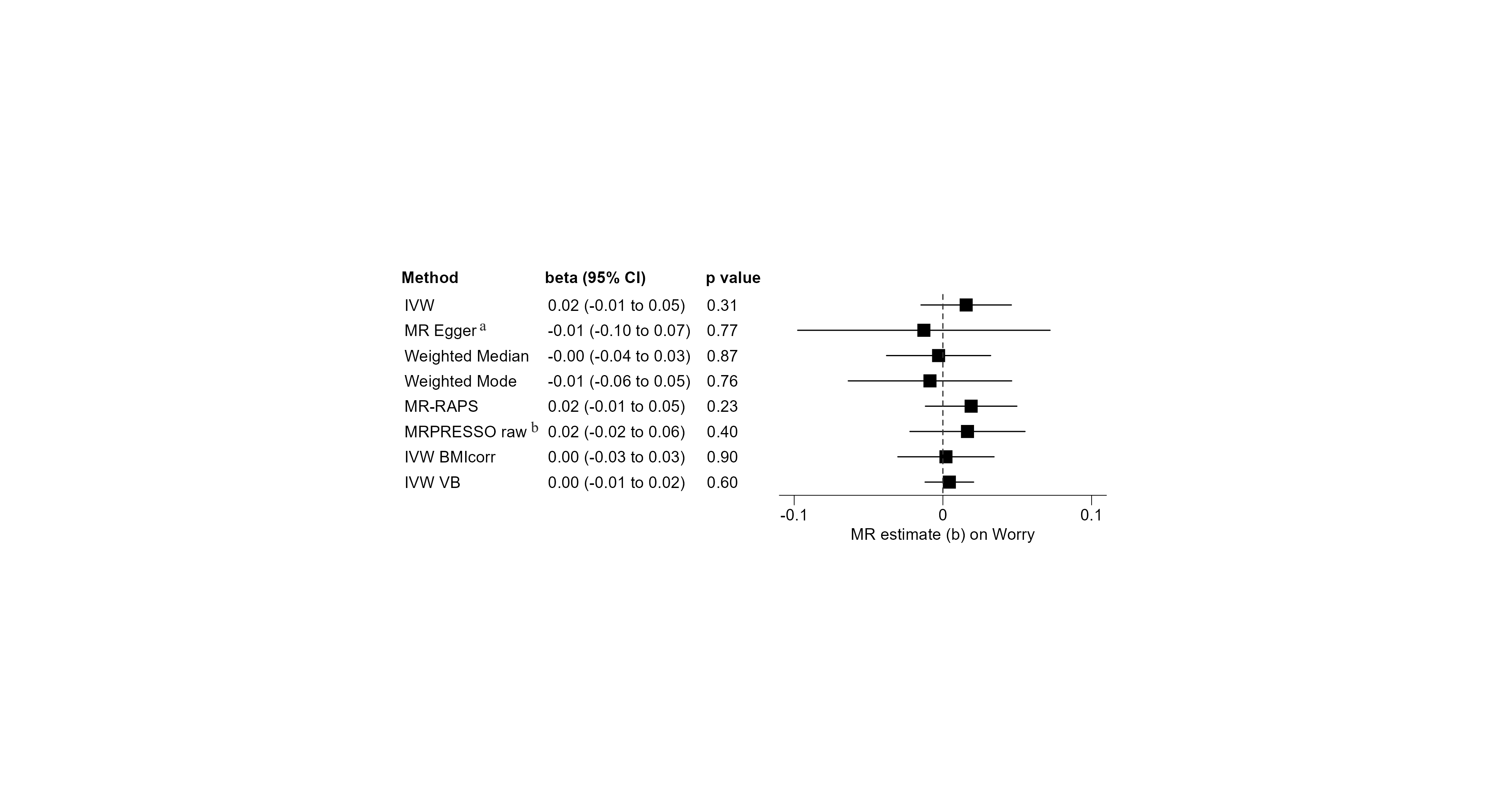

**Supplementary Figure S30 – Sensitivity Analyses – Worry.** Univariable MR analyses on the effect of male puberty timing on the ‘Worry’ subdomain of neuroticism. 72 of the 76 target SNPs for male puberty timing were covered in the outcome GWAS by Nagel, et al. [18]. Positive MR estimates indicate that later male puberty timing is associated with higher expression on the ‘Worry’ subdomain of neuroticism. The Q-statistic did reveal significant heterogeneity (IVW: df=66, Q-statistic: 118.80, p=7.3x10^-5^; MR Egger: df=65, Q-statistic: 117.90, p=6.6x10^-5^). a: Eggers-intercept did not indicate significant directional pleiotropy (intercept=0.0009, se=0.0012, p=0.484). The I^2^-statistic was 0.792, and thus below the threshold of 0.9, indicating a violation of the NOME assumption. Therefore, the MR Egger result should be interpreted with caution. b: The MR-PRESSO global test for pleiotropy was significant (RSSobs=224.89, p<3x10^-4^), and five outliers were identified by MR-PRESSO (see Supplemental Table S2 for details). Thus, the instrumental variable consisted of 67 SNPs.

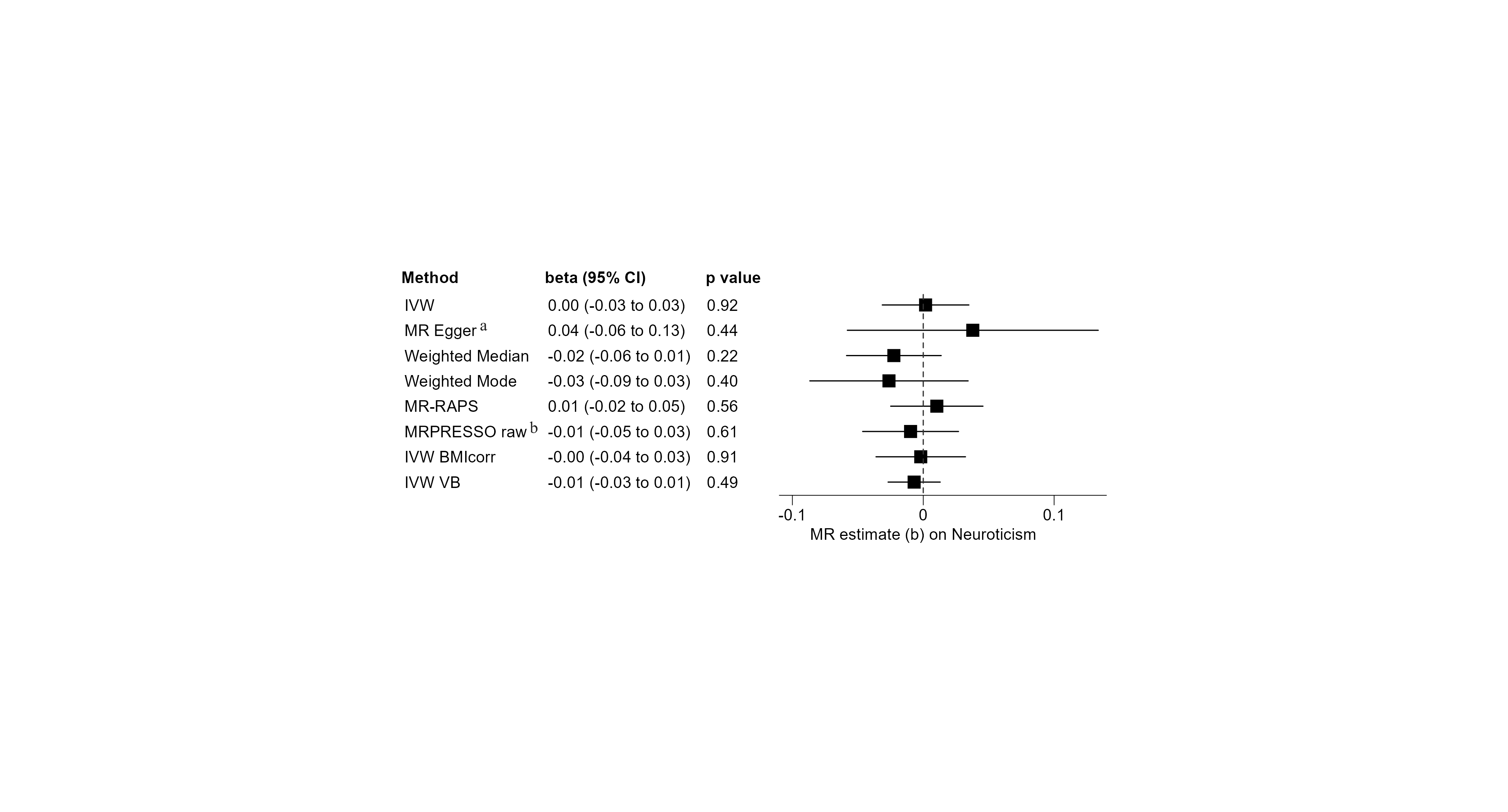

**Supplementary Figure S31 – Sensitivity Analyses – Neuroticism.** Univariable MR analyses on the effect of male puberty timing on neuroticism. 72 of the 76 target SNPs for male puberty timing were covered in the outcome GWAS by Nagel, et al. [18]. Positive MR estimates indicate that later male puberty timing is associated with higher expression of neuroticism. The Q-statistic did reveal significant heterogeneity (IVW: df=68, Q-statistic: 152.11, p= 2.2x10^-8^; MR Egger: df=67, Q-statistic: 150.72, p=2.2x10^-8^). a: Eggers-intercept did not indicate significant directional pleiotropy (intercept=-0.0011, se=0.0014, p=0.434). The I^2^-statistic was 0.732, indicating a relevant violation of the NOME assumption. b: The MR-PRESSO global test for pleiotropy was significant (RSSobs=218.53, p<3x10^-4^), and three outliers were identified by MR-PRESSO (see Supplemental Table S2 for details). Thus, the instrumental variable consisted of 69 SNPs.

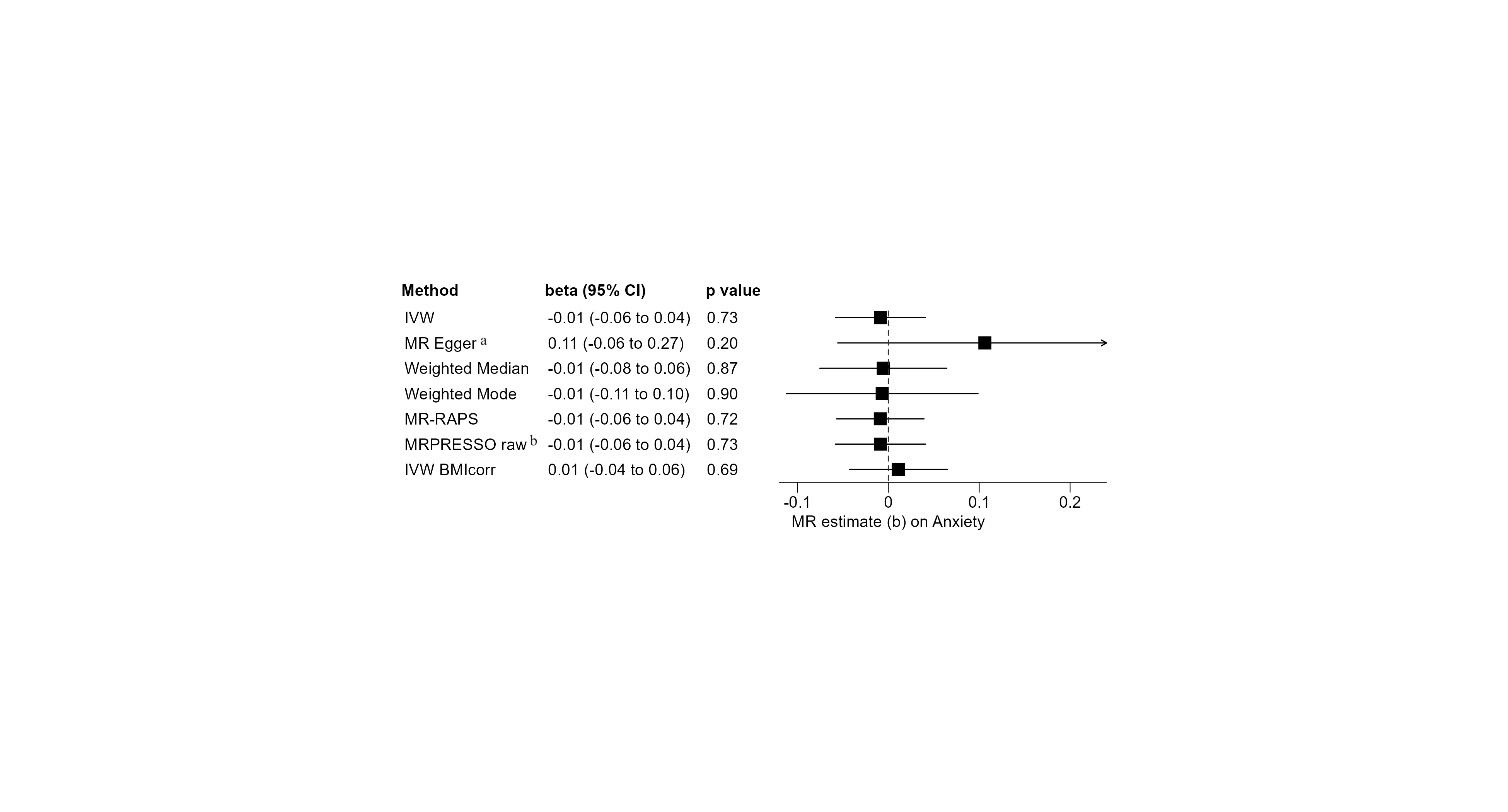

**Supplementary Figure S32 – Sensitivity Analyses – Anxiety Disorders.** Univariable MR analyses on the effect of male puberty timing on the development and/or severity of anxiety disorders (factor score). 73 of the 76 target SNPs for male puberty timing were covered in the outcome GWAS by Otowa, et al. [23]. Positive MR estimates indicate that later male puberty timing is associated with a higher risk for and/or higher disease severity of anxiety disorders. The Q-statistic did not reveal significant heterogeneity (IVW: df=72, Q-statistic: 79.22, p=0.262; MR Egger: df=71, Q-statistic: 76.91, p= 0.295). a: Eggers-intercept did not indicate significant directional pleiotropy (intercept=-0.0034, se=0.0023, p=0.149). The I^2^-statistic was 0.937, indicating no major violation of the NOME assumption. b: The MR-PRESSO global test for pleiotropy was not significant (RSSobs=80.90, p=0.278), and no outlier was identified by MR-PRESSO (see Supplemental Table S2 for details). Thus, the MR-PRESSO corrected IVW estimate (IVW) corresponds to the uncorrected IVW estimate (MR-PRESSO raw).

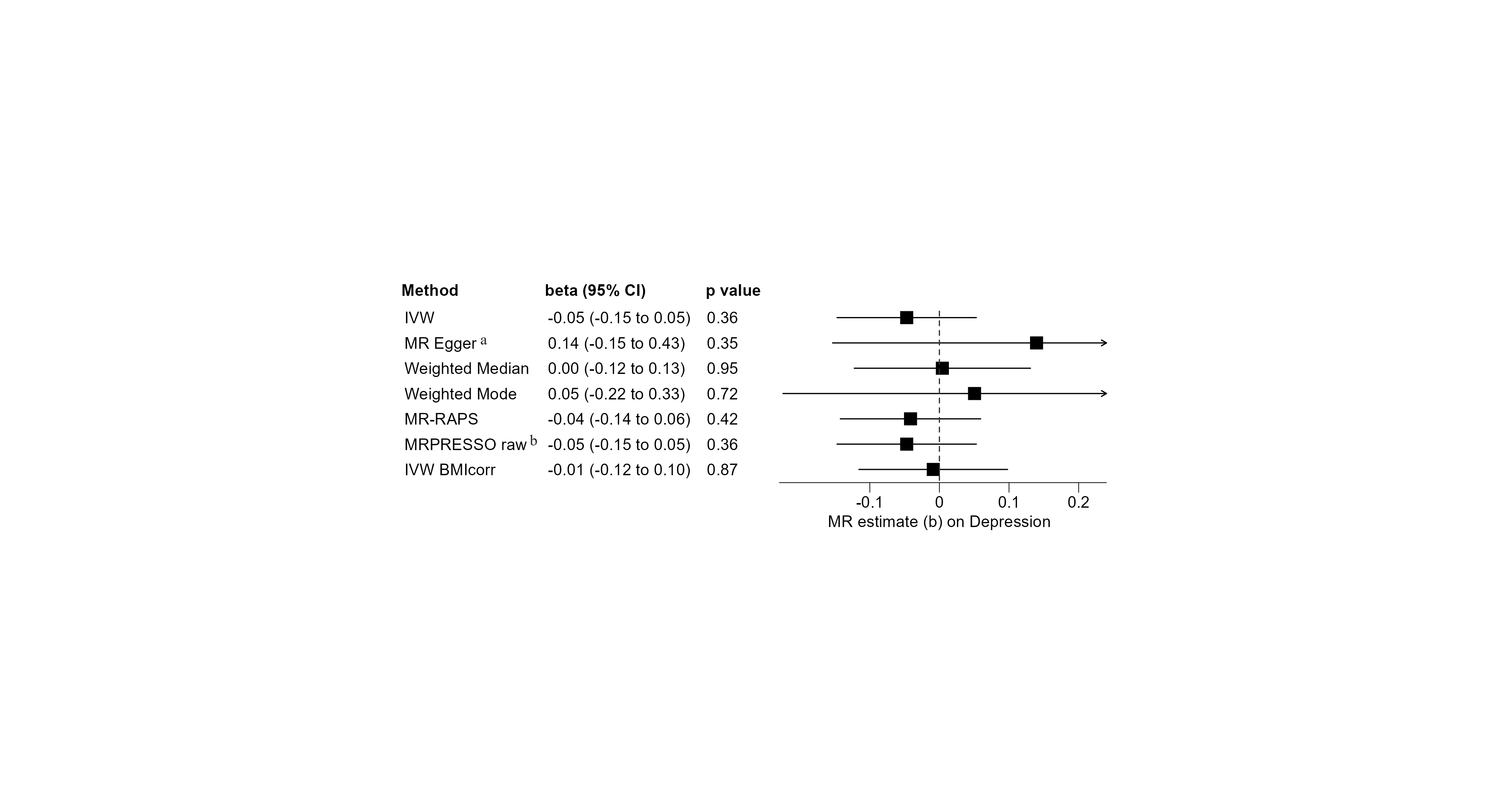

**Supplementary Figure S33 – Sensitivity Analyses – Depression.** Univariable MR analyses on the effect of male puberty timing on the risk of developing a major depressive disorder. 76 of the 76 target SNPs for male puberty timing were covered in the outcome GWAS by Wray, et al. [21]. Positive MR estimates indicate that later male puberty timing is associated with a higher risk for developing depression. The Q-statistic did reveal significant heterogeneity (IVW: df=75, Q-statistic: 110.26, p=0.005; MR Egger: df=74, Q-statistic: 107.70, p=0.006). a: Eggers-intercept did not indicate significant directional pleiotropy (intercept=-0.0056, se=0.0042, p=0.189). The I^2^-statistic was 0.776, and thus below the threshold of 0.9, indicating a violation of the NOME assumption. Therefore, the MR Egger result should be interpreted with caution. b: The MR-PRESSO global test for pleiotropy was significant (RSSobs=113.16, p=0.006). However, no significant outlier was identified by MR-PRESSO (see Supplemental Table S2 for details). Thus, the MR-PRESSO corrected IVW estimate (IVW) corresponds to the uncorrected IVW estimate (MR-PRESSO raw).
